# Mitochondrial DNA haplogroups and trajectories of cardiometabolic risk factors during childhood and adolescence: a prospective cohort study

**DOI:** 10.1101/2020.11.06.20226985

**Authors:** Kate N O’Neill, Emily Aubrey, Laura D Howe, Evie Stergiakouli, Santiago Rodriguez, Patricia M Kearney, Linda M O’Keeffe

**Author notes:** **Corresponding author:** Dr Kate O’Neill, School of Public Health, 4^th^ Floor Western Gateway Building, University College Cork, Ireland. **Disclosures:** None of the authors have any conflicts of interest to declare.

## Abstract

**Background:** Mitochondria are organelles responsible for converting glucose into energy. Mitochondrial DNA is exclusively maternally inherited by offspring. The role of mitochondrial DNA haplogroups in the aetiology of cardiometabolic disease risk is not well understood.

**Methods:** We examined the sex-specific association between European mitochondrial DNA haplogroups and trajectories of nine cardiometabolic risk factors from birth to 18 years in a prospective English birth cohort. Mitochondrial haplogroups were analysed according to common European haplogroups; H,U,J,T,K,V,W,I and X. Nine cardiometabolic risk factors measured over varying times from birth/mid-childhood to age 18 years included body mass index (BMI), fat mass and lean mass, systolic blood pressure (SBP), diastolic blood pressure (DBP), pulse rate, high-density lipoprotein cholesterol (HDL-c), non HDL-c and triglycerides. Fractional polynomial and linear spline multilevel models stratified by sex explored the sex-specific association between haplogroups and risk factor trajectories.

**Results:** Among 6,360-7,954 participants with 22,864-79,178 repeated measures per outcome, we found no strong evidence that haplogroups U,T,J,K and W were associated with trajectories of cardiometabolic risk factors across childhood and adolescence compared to haplogroup H. In females, haplogroup V was associated with 4.0% (95% CI: 1.4, 6.7) lower BMI at age 7 years and 9.3% (95% CI: 1.9, 16.7) lower fat mass at age 9, though differences did not persist at age 18. Haplogroup X was associated with 1.3kg (95% CI: 0.5, 2.2) lower lean mass and 16.4% (95% CI: 3.5, 29.3) lower fat mass at age 9; associations with lower lean mass persisted at 18 years whereas associations with fat mass did not. In males, haplogroup I was associated with 2.4% (95% CI: 0.2, 4.6) higher BMI at age 7; this difference widened to 5.1% (95% CI: 0.9,9.3) at 18 years.

**Conclusion:** Our study demonstrated some evidence of sex-specific associations between mitochondrial DNA haplogroups V, I and X and trajectories of adiposity during childhood and adolescence.

## Background

Mitochondria are organelles responsible for converting glucose into adenosine triphosphate, a form of energy that our cells are able to use (1,2). This process requires proteins, a small number of which are coded for by circular mitochondrial deoxyribose nucleic acid (DNA). Unlike nuclear DNA, mitochondrial DNA does not recombine and is exclusively maternally inherited by offspring (3). Mutations throughout human history resulted in subdivisions of mitochondrial DNA into discrete region-specific haplogroups (4). For instance, in Europe, 90% of the population belong to one of five major haplogroups (5). Variation in mitochondrial DNA may play a role in the aetiology of numerous diseases (6), including cardiometabolic disease (2) and cancer (7) as well as longevity (8), through differential production of reactive oxygen species that may impact oxidative stress of cells (2,3,8).

Evidence to date on the association between mitochondrial DNA haplogroups and cardiometabolic risk is conflicting (9–12). In a case-control study of 3,889 participants in the UK, haplogroup K was associated with risk of ischemic vascular events (12). In a prospective cohort study of 3,288 participants in the USA and a case-control study of 781 participants in Spain, haplogroup J was associated with lower risk of ischemic cardiomyopathy and cardiovascular disease (CVD) (9,10). In contrast, a large prospective population-based study of 9,254 participants in Denmark did not find any strong evidence of associations between common European haplogroups and risk of ischaemic CVD (11). Evidence has also been inconsistent for associations with key cardiometabolic risk factors. For instance, haplogroup X was associated with lower body mass index (BMI) and fat mass in a cross-sectional study of 2,286 adults in the US and haplogroup T was associated with higher risk of obesity in a case-control study of 716 adults in Southern Italy (13,14). In contrast, a case-control of 4,070 adults in Germany did not find evidence of associations between mitochondrial DNA haplogroups and obesity (15). In addition, most studies have examined associations in adults only (9–14,16), despite the early life origins of cardiometabolic risk which track through the life course (17,18). Furthermore, studies to date have not examined the sex-specific association of mitochondrial DNA with cardiometabolic risk factors (9–16,19) although there are established sex differences in cardiometabolic risk across the life course (20–23).

Using data from a contemporary prospective birth cohort study in the Southwest of England, we examined the sex-specific associations between common European mitochondrial DNA haplogroups and cardiometabolic risk factors during childhood and adolescence. Risk factors included BMI (one to 18 years), height-adjusted fat and lean mass (9 to 18 years), systolic blood pressure (SBP), diastolic blood pressure (DBP) and pulse rate (7 to 18 years), high density lipoprotein cholesterol (HDL-c), non-high density lipoprotein cholesterol (non-HDL-c) and triglycerides (birth to 18 years).

## Methods

### Study participants

The Avon Longitudinal Study of Parents and Children (ALSPAC) is a prospective birth cohort study in Southwest England (24,25). Pregnant women resident in Avon, UK with expected dates of delivery 1st April 1991 to 31st December 1992 were invited to take part in the study. The initial number of pregnancies enrolled is 14,541 (for these at least one questionnaire has been returned or a “Children in Focus” clinic had been attended by 19/07/99). Of these initial pregnancies, there was a total of 14,676 foetuses, resulting in 14,062 live births and 13,988 children who were alive at 1 year of age. When the oldest children were approximately 7 years of age, an attempt was made to bolster the initial sample with eligible cases who had failed to join the study originally. Therefore, the total sample size for analyses using any data collected after the age of 7 is 15,454 pregnancies, resulting in 15,589 foetuses. Of these 14,901 were alive at 1 year of age. The study has been described elsewhere in detail (24,25). Follow-up includes parent and child completed questionnaires, links to routine data and clinic attendance. Research clinics were held when the children were aged approximately 7, 9, 10, 11, 13, 15 and 18 years old. Ethical approval for the study was obtained from the ALSPAC Ethics and Law Committee and the Local Research Ethics Committees. The study website contains details of all the data that is available through a fully searchable data dictionary http://www.bristol.ac.uk/alspac/researchers/our-data/.

### Mitochondrial DNA haplogroup derivation

Children had DNA sampled at either birth from cord blood or at a research clinic at age 7. A total of 9,912 children were genotyped using the SNP genotyping platform by Sample Logistics and Genotyping Facilities at the Wellcome Trust Sanger Institute and LabCorp (Laboratory Corporation of America) using support from 23andMe. After quality control assessment methods and removing individuals who withdrew consent; our dataset contained 8,209 individuals with derived mitochondrial DNA haplogroups. Our analysis included nine common European haplogroup categories, H, U, T, J, K, V, I, W and X. Individuals with rare European and non-European haplogroups were excluded (A, C, D, L, M, N, R; n=107). Further information on how haplogroups were derived including the frequency of each haplogroup are included in Appendix 1 and Supplementary Table S1. Figure S1 shows a flow diagram of participants eligible for inclusion.

### Cardiometabolic risk factor measurement

#### BMI, Fat and Lean Mass

Length (before the age of 2 years), height (from the age of 2 years) and weight data from the age of 1 year were obtained from several sources including health visitor records, questionnaires and clinics from birth to 18 years. BMI was calculated as weight (kg) divided by height squared (m^2^). Whole body less head, and central fat and lean mass were derived from whole body dual energy X-ray absorptiometry (DXA) scans assessed 5 times at ages 9, 11, 13, 15, and 18 using a Lunar prodigy narrow fan beam densitometer.

#### SBP, DBP and pulse rate

At each clinic (ages 7, 9, 10, 11, 12, 15 and 18), SBP, DBP and pulse rate were measured at least twice in each with the child sitting and at rest with the arm supported, using a cuff size appropriate for the child’s upper arm circumference and a validated blood pressure monitor. The mean of the two final measures was used.

#### Blood based biomarkers

HDL-c, total cholesterol and triglycerides were measured in cord blood at birth and from venous blood subsequently. Samples were non-fasted at 7 and 9 years; fasting measures were available from clinics at 15 and 18 years. Non-HDL-c was calculated by subtracting HDL-c from total cholesterol at each measurement occasion. Trajectories of HDL-c, non-HDL-c and triglycerides were derived from a combination of measures from cord blood, non-fasting and fasting bloods.

Further information on details of measurement sources are included in Appendix 2 of Supplementary Material.

### Statistical analysis

We used multilevel models to examine the association between mitochondrial DNA haplogroups and change in each risk factor across childhood and into adolescence (26,27). Multilevel models estimate mean trajectories of the risk factor while accounting for the non-independence (i.e. clustering) of repeated measurements within individuals, change in scale and variance of measures over time, and differences in the number and timing of measurements between individuals (28,29). We included all participants with at least one measure of the risk factor in each multilevel model, under a missing-at-random (MAR) assumption, to minimise selection bias (28,29). Sex-specific trajectories of all outcomes have been modelled previously using multilevel models and are described elsewhere in detail (20,30– 32). Trajectories of BMI were modelled using fractional polynomials while trajectories of all other risk factors (fat mass, lean mass, SBP, DBP, pulse rate, HDL-c, non-HDL-c, triglycerides) were estimated using linear spline multilevel models (all models had two levels: measurement occasion and individual). Further information on the modelling of these trajectories including how fractional polynomial terms and spline periods were selected are detailed in Appendix 3 of Supplementary Material. Model fit statistics for each risk factor trajectory are shown in Supplementary Material Tables S2-S8.

### Association between mitochondrial DNA haplogroups and trajectories

Participants with data on sex, mitochondrial DNA haplogroup and at least one measure of a risk factor were included in analyses. Those who reported being pregnant at the 18-year clinic (n=6) were excluded from analyses at that time point only. To explore the sex-specific association of mitochondrial DNA haplogroups with the above sex-specific trajectories of cardiometabolic risk factors, males and females were analysed separately. In all analyses, haplogroup H as the most common in the ALSPAC sample and in European populations was selected as the reference haplogroup. An interaction between each of the other haplogroups (U,T,J,K,V,W,I,X) and both the intercept and each fractional polynomial term (BMI) or spline (all other risk factors) was included in the models to estimate the difference in intercepts and slopes of each cardiometabolic risk factor between these haplogroups and the reference haplogroup, haplogroup H. All trajectories were modelled in MLwiN version 3.04 (33), called from Stata version 16 (34) using the runmlwin command (35).

In all models, age (in years) was centred at the first available measure. Values of cardiometabolic risk factors that had a skewed distribution (BMI, fat mass, triglycerides) were (natural) log transformed prior to analyses. Differences and confidence intervals for these risk factors were calculated on the log-scale and then back-transformed and converted to percentage change for simplicity of interpretation.

## Results

Table 1 shows characteristics of participants by sex. Maternal education, household social class and maternal smoking during pregnancy were similar in males and females. Table 1 and Table S9 shows the frequency of each haplogroup analysed in the study. Haplogroup H was the most frequent in the population, representing approximately 45% of males and females. The number of participants included in the analyses ranged from a total of 3,216 females (11,909 measures) and 3,144 males (10,955 measures) for fat mass, up to 3,886 females and 4,068 males for BMI (total measures of 39,778 and 39,400 respectively) (Table S10, Figure S1). Compared with participants excluded from analyses, those included tended to have higher maternal education, higher household social class and lower maternal smoking during pregnancy (Table S11).

**Table 1.**
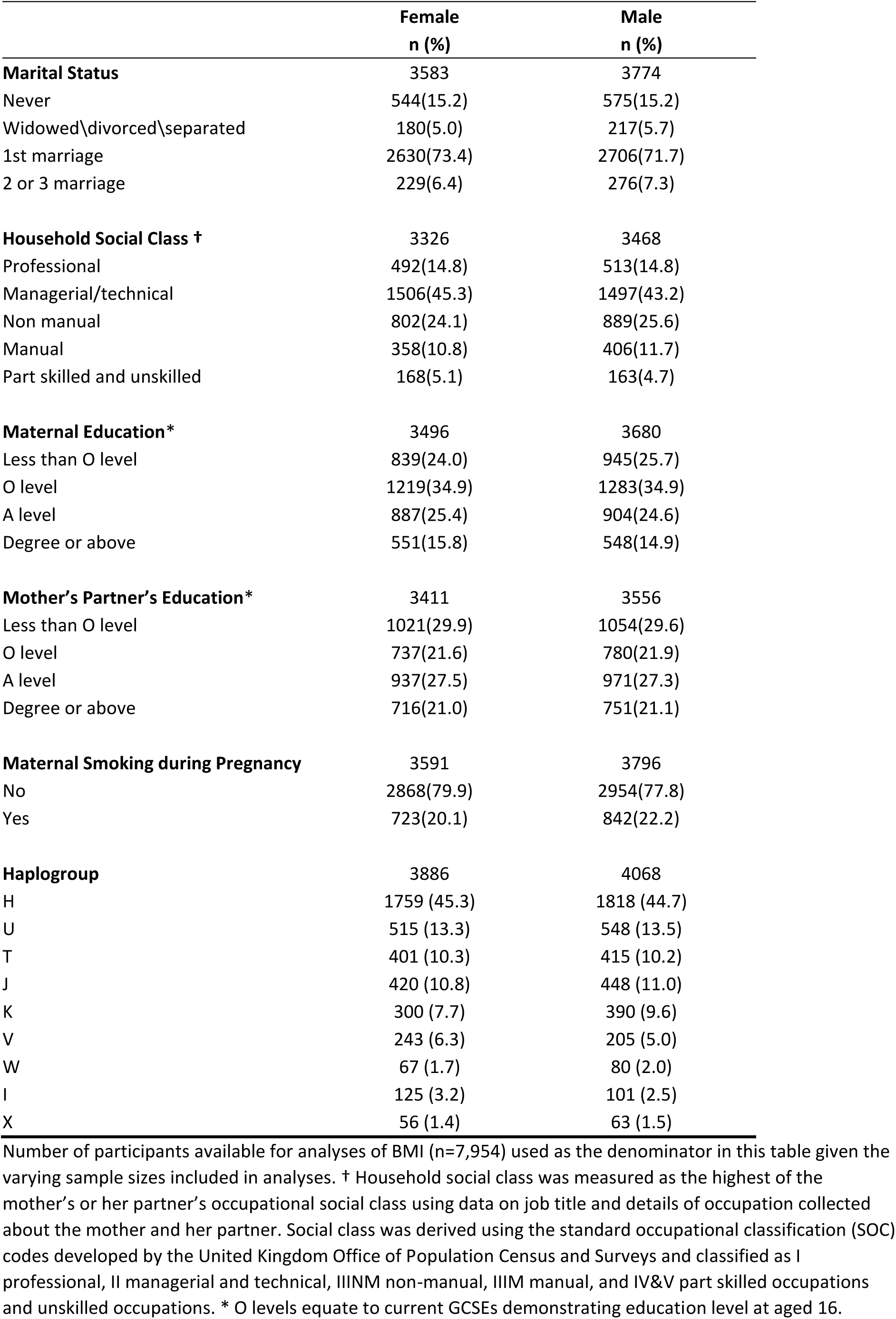
Characteristics of ALSPAC participants included in analysis, by sex.

|  | **Female**<br>**n (%)** | **Male**<br>**n (%)** |
| --- | --- | --- |
| **Marital Status** | 3583 | 3774 |
| Never | 544(15.2) | 575(15.2) |
| Widowed\divorced\separated | 180(5.0) | 217(5.7) |
| 1st marriage | 2630(73.4) | 2706(71.7) |
| 2 or 3 marriage | 229(6.4) | 276(7.3) |
| **Household Social Class †** | 3326 | 3468 |
| Professional | 492(14.8) | 513(14.8) |
| Managerial/technical | 1506(45.3) | 1497(43.2) |
| Non manual | 802(24.1) | 889(25.6) |
| Manual | 358(10.8) | 406(11.7) |
| Part skilled and unskilled | 168(5.1) | 163(4.7) |
| **Maternal Education\*** | 3496 | 3680 |
| Less than O level | 839(24.0) | 945(25.7) |
| O level | 1219(34.9) | 1283(34.9) |
| A level | 887(25.4) | 904(24.6) |
| Degree or above | 551(15.8) | 548(14.9) |
| **Mother's Partner's Education\*** | 3411 | 3556 |
| Less than O level | 1021(29.9) | 1054(29.6) |
| O level | 737(21.6) | 780(21.9) |
| A level | 937(27.5) | 971(27.3) |
| Degree or above | 716(21.0) | 751(21.1) |
| **Maternal Smoking during Pregnancy** | 3591 | 3796 |
| No | 2868(79.9) | 2954(77.8) |
| Yes | 723(20.1) | 842(22.2) |
| **Haplogroup** | 3886 | 4068 |
| H | 1759 (45.3) | 1818 (44.7) |
| U | 515 (13.3) | 548 (13.5) |
| T | 401 (10.3) | 415 (10.2) |
| J | 420 (10.8) | 448 (11.0) |
| K | 300 (7.7) | 390 (9.6) |
| V | 243 (6.3) | 205 (5.0) |
| W | 67 (1.7) | 80 (2.0) |
| I | 125 (3.2) | 101 (2.5) |
| X | 56 (1.4) | 63 (1.5) |

Number of participants available for analyses of BMI (n=7,954) used as the denominator in this table given the varying sample sizes included in analyses. † Household social class was measured as the highest of the mother's or her partner's occupational social class using data on job title and details of occupation collected about the mother and her partner. Social class was derived using the standard occupational classification (SOC) codes developed by the United Kingdom Office of Population Census and Surveys and classified as I professional, II managerial and technical, IIIM non-manual, IIIM manual, and IV&V part skilled occupations and unskilled occupations. \* O levels equate to current GCSEs demonstrating education level at aged 16.

### BMI, Fat and Lean Mass

Among females, haplogroups U,T,J,K and I were not strongly associated with trajectories of BMI from one to 18 years and trajectories of fat mass and lean mass from 9 to 18 years. (Figure 1, Table S12). Haplogroup V was associated with 4.0% (95% confidence interval (CI): −6.7, −1.4) lower BMI at age 3 years, though this difference did not persist at age 18 (difference; −1.3%; 95% CI: −3.9, 1.4). Haplogroup V was associated with 9.3% (95% CI: −16.7, −1.9) lower fat mass at age 9 years; this difference reversed over childhood and adolescence, such that by age 18, haplogroup V was associated with −1.0 kg (95% CI: −1.7, −0.3) lower lean mass. Haplogroup X was associated with 16.4% (95% CI: 3.5, 29.3) lower fat mass at age 9 but this difference did not persist at age 18 (difference; 8.9% (95% CI: −12.8, 30.6)) (Figure 1, Table S13). Haplogroup W and X were associated with lower lean mass at age 9; this association persisted at age 18 for haplogroup X (difference; −1.4kg, 95% CI: −2.8, −0.02) but not for haplogroup W (difference; 0.04kg, 95% CI: −1.3, 1.4) (Figure 1, Table S14).

**Figure 1:**
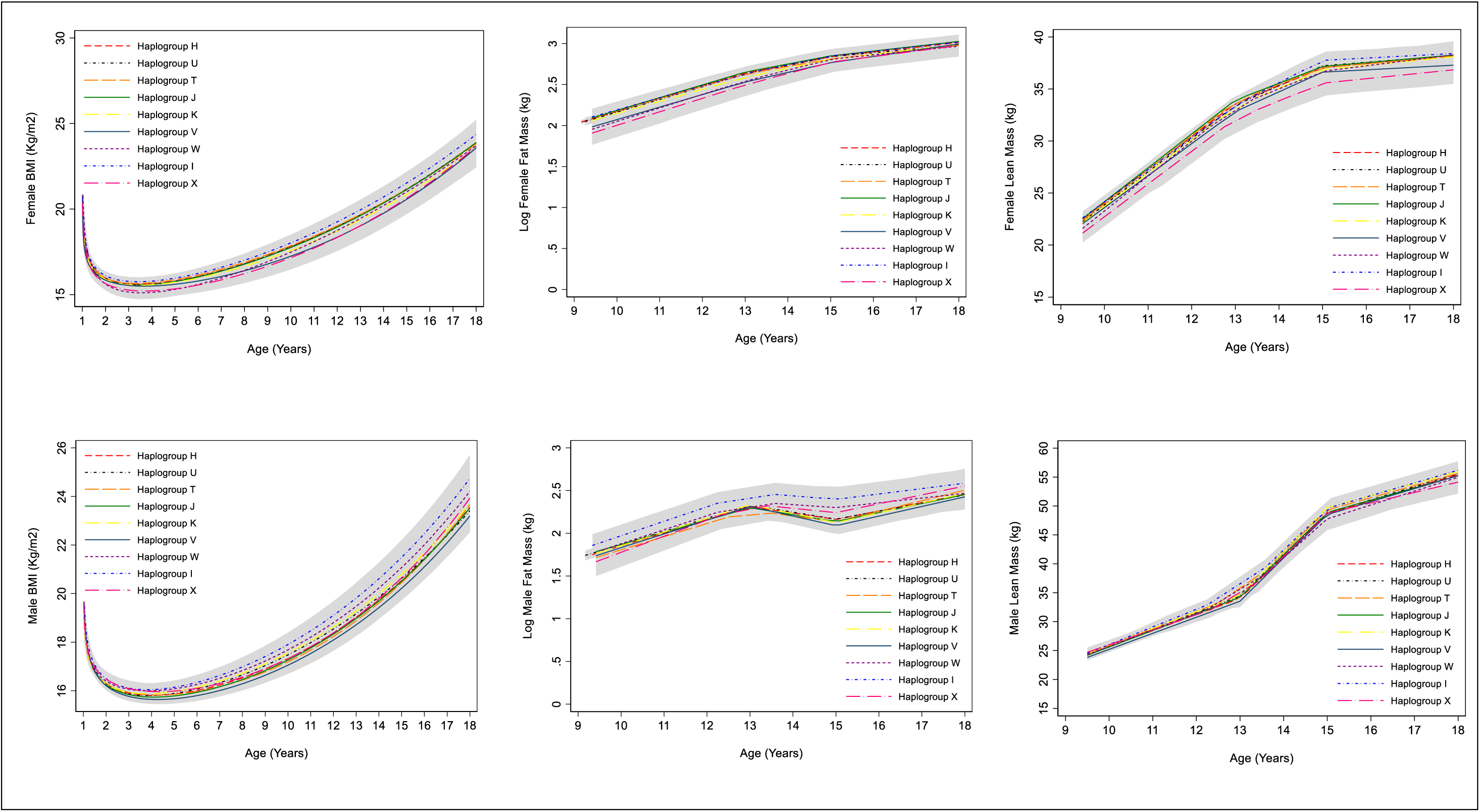
Mean trajectories of BMI, log fat mass and lean mass in females and males, by haplogroup. 95% confidence intervals for all haplogroups are displayed in grey. Detailed results with confidence intervals are provided in Supplementary Tables S12-14. Note the different age range on the X axis for each outcome. BMI, body mass index.

Among males, haplogroup I was associated with 2.4% (95% CI: 0.2, 4.6) higher BMI at 7 years; this increased to a difference of 5.1% (95%CI: 0.9, 9.3) at 18 years. Haplogroup I was also associated with higher fat mass in males at age 18 years only, albeit with wide confidence intervals (difference; 31.8% (95% CI: 0.3, 63.4)). Haplogroup X was associated with higher fat mass at age 18, although with confidence intervals spanning the null (37.7%, 95% CI: −4.2, 79.7).

### SBP, DBP, Pulse

We found no strong evidence of associations between any haplogroup and trajectories of SBP, DBP or pulse from ages 7 to 18 years in females and males (Figure 2, Table S15).

**Figure 2:**
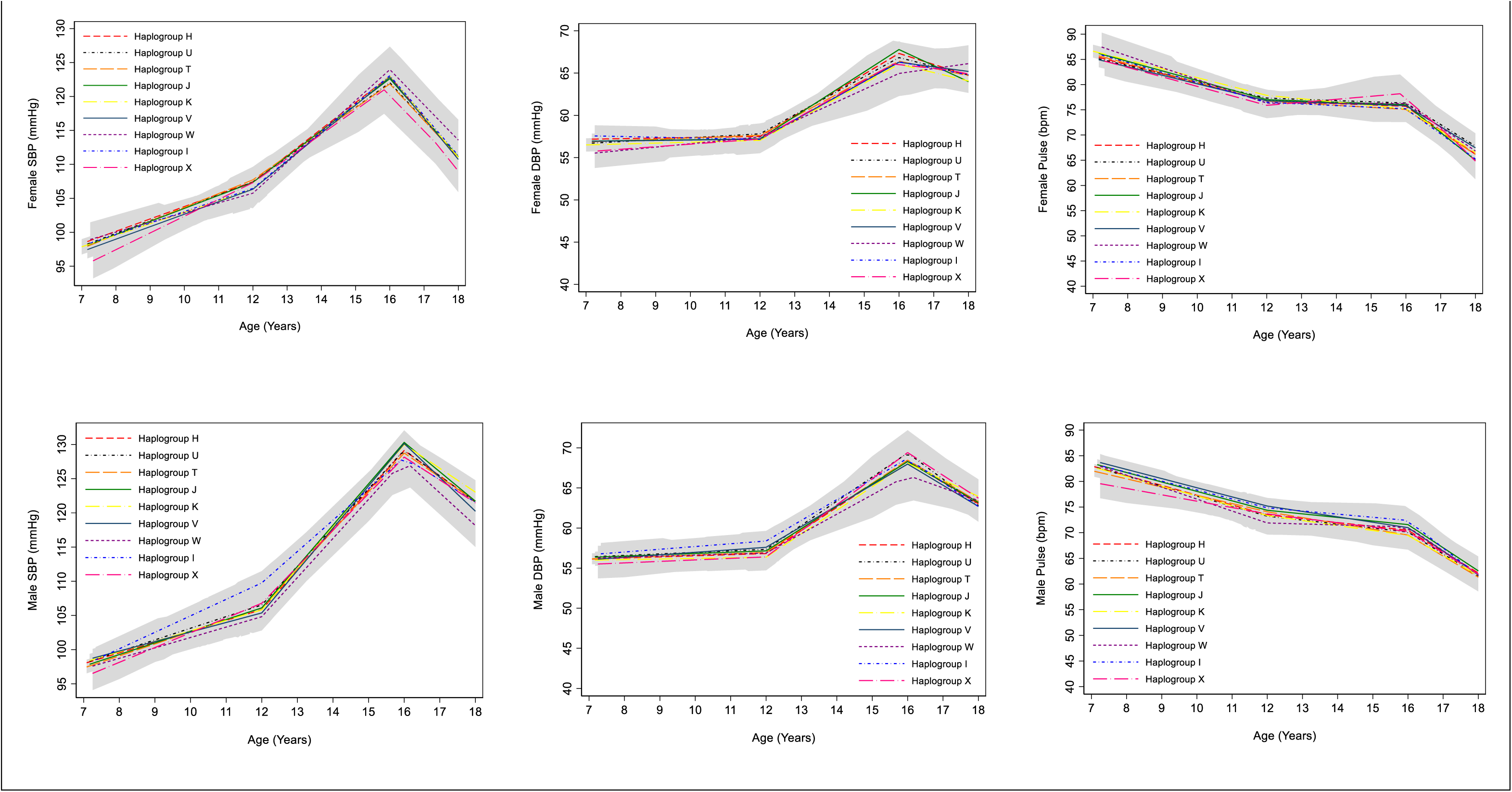
Mean trajectories of SBP, DBP and pulse in females and males, by haplogroup. 95% confidence intervals for all haplogroups are displayed in grey. Detailed results with confidence intervals are provided in Supplementary Table S15. SBP, systolic blood pressure; DBP, diastolic blood pressure.

### Blood based biomarkers

We found no strong evidence of associations between haplogroups U,T,J,K,V,I and X and trajectories of HDL-c and non-HDL-c from birth to 18 years in females and males (Figure 3, Table S16). Haplogroup W was associated with 2.1 mmol/l (95% CI: 1.1, 3.2) higher HDL-c at birth in males and 1.5mmol/l (95% CI: 0.7, 2.4) lower non-HDL-c but this difference did not persist at age 18 years (HDL difference: 0.05mmol/l (95% CI: −0.03, 0.1); non-HDL difference: 0.2mmol/l (95% CI: −0.1,0.4)). We found no strong evidence of associations between haplogroups and triglyceride trajectories of females and males from birth to 18 years (Figure 3, Table S17).

**Figure 3:**
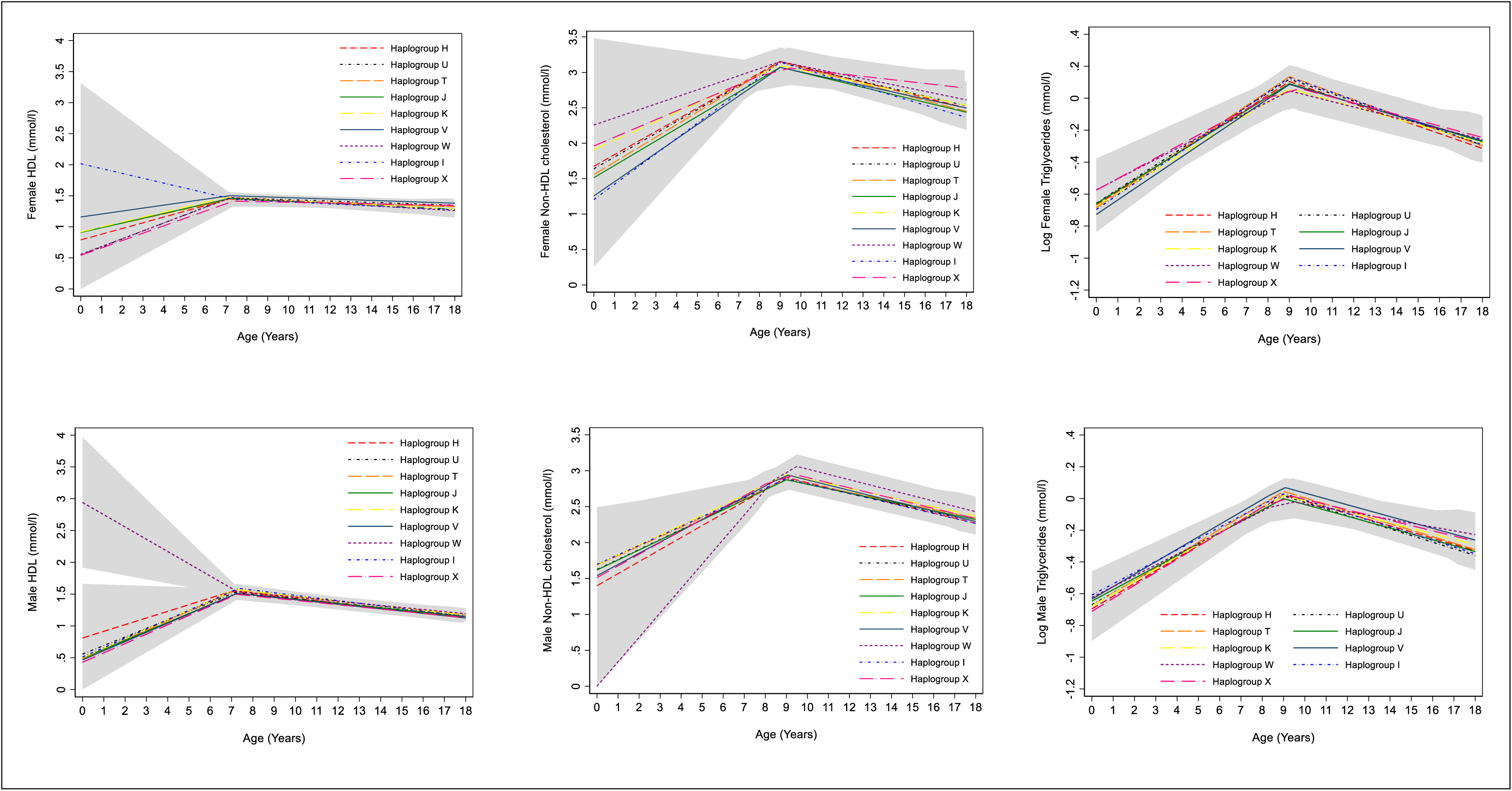
Mean trajectories of HDL-c, Non-HDL-c and log triglycerides in females and males, by haplogroup. 95% confidence intervals for all haplogroups are displayed in grey. Detailed results with confidence intervals are provided in Supplementary Tables S16 & S17. Note the different age range on the X axis for each outcome. HDL-c, high density lipoprotein cholesterol.

## Discussion

In this large prospective cohort study, we examined the sex-specific association between mitochondrial DNA haplogroups and longitudinal changes in nine cardiometabolic risk factors from birth or early childhood through to 18 years. We found some evidence of sex-specific associations of haplogroup V with BMI, fat mass and lean mass in females, haplogroup X with fat mass and lean mass in females and haplogroup I with BMI and fat mass in males.

### Comparison with other studies

Most studies to date have examined the association between mitochondrial DNA and cardiometabolic risk factors in adults and have not explored the contribution of mitochondrial DNA to the sex-specific aetiology of cardiometabolic risk across the early life course (9,11,16). However, our findings are comparable with some previous analyses of females and males combined. For example, we found that haplogroup I among males was associated with higher BMI at age 7 that persisted at age 18 years and higher fat mass at 18 years. This is consistent with findings in a prospective study of 2,342 participants aged 45 to 79 years recruited from four clinical sites in the US which showed increased incidence of obesity in males and females with haplogroup I, W and X combined (16). Our findings build on this evidence, indicating that the observed increased BMI associated with haplogroup I, X and W combined in that study may be driven predominantly by haplogroup I, is potentially driven by fat mass rather than lean mass, may be unique to males and emerge in childhood, potentially tracking through to adulthood. Similarly, our findings of lower fat mass at age 9 and lower lean mass from age 9 to 18 years in females with haplogroup X are consistent with previous findings in a sex-combined analysis of 2,286 Caucasian adults of Northern European origin living in the US with a mean age of 51 years (14) which found lower BMI and fat mass in adults with haplogroup X. Our results indicate that associations of haplogroup X with BMI may be specific to females, are potentially driven by associations with both fat and lean mass and potentially begin in early life.

Our findings contrast with a previous population-based study of 9,254 adults in Denmark (11) which did not find evidence of associations between mitochondrial DNA haplogroups and cardiometabolic risk factors. Our results also contrast with two case-control studies comparing those with obesity to those without (15,36), which found no evidence of associations of obesity with any haplogroup. Furthermore, we also found no strong evidence of an association of haplogroup T and BMI or fat mass, as found in two additional case-control studies, one conducted in Southern Italy with 716 adults (13) and another Austrian study with 514 juveniles (aged less than 21 years) and 1,598 adults (19). However, we did find evidence of associations of haplogroup V and lower BMI, lower fat mass at age 9 and lower lean mass at age 18 in females and evidence of an association between haplogroup W and HDL-c at birth in males which to our knowledge has not been demonstrated previously. Reasons for differences in findings between our work and previous studies may include the examination of mitochondrial DNA haplogroups and outcomes in both sexes combined in previous studies, thus potentially overlooking the sex-specific aetiology of mitochondrial DNA haplogroups and cardiometabolic risk examined here. A further possibility for differences between our study and previous work may include our examination of trajectories of repeated continuous measures of risk factors from birth/early childhood to 18 years, allowing us to examine when associations emerge and whether they persist over time in females and males. Further work is required to extend such analyses beyond 18 years to examine if mitochondrial DNA haplogroups are associated with changes in risk factors from early and mid-adulthood into older age.

### Strengths and limitations

There are a number of strengths to our study including the availability of repeated measures of cardiometabolic risk factors throughout from birth/early to age 18 years and the use of multilevel models allowing for clustering of repeated measures within individuals and correlation between measures over time. Our study is the first to explore the association between mitochondrial DNA haplogroups and change over time in cardiometabolic risk factors from childhood to adolescence. Due to the large sample size, we were able to examine rarer haplogroups such as W, I and X separately. In addition, we have conducted sex-specific analyses to account for the differences in risk factor trajectories previously reported (20) and to better understand the associations between mitochondrial DNA haplogroups and cardiometabolic risk factors in females and males. The study also has some limitations. Due to sparsity of measures we were only able to explore linear change over time. Loss-to-follow up is another potential limitation of our study. However, we have included all participants with at least one measure of each risk factor to minimise any potential selection bias. The number of people with measurements of each risk factor also varied meaning that our sample sizes differed for each risk factor and thus are not directly comparable. Our sample may be underpowered to detect effects for rarer haplogroups and additionally some observed associations may be a result of type 1 error due to multiple testing.

## Conclusion

Our study demonstrated some evidence of sex-specific associations between mitochondrial DNA haplogroups V, I and X and trajectories of adiposity during childhood and adolescence.

## Data Availability

Data are available upon submission and approval of a research proposal to the ALSPAC Executive.
Further information can be found at http://www.bristol.ac.uk/alspac/researchers/access/

## Acknowledgements

We are extremely grateful to all the families who took part in this study, the midwives for their help in recruiting them, and the whole ALSPAC team, which includes interviewers, computer and laboratory technicians, clerical workers, research scientists, volunteers, managers, receptionists and nurses.

