## Supplementary Material for "Mitochondrial DNA haplogroups and trajectories of cardiometabolic risk factors during childhood and adolescence: a prospective cohort study"

Kate N O’Neill ^1^, PhD; Emily Aubrey ^2, 3^ MB ChB; Laura D Howe ^2, 3^, PhD; Evie Stergiakouli ^2,3^, PhD; Santiago Rodriguez ^2,3^, PhD; Patricia M Kearney ^1^ , PhD; Linda M O’Keeffe ^1, 2, 3^, PhD

^1^ School of Public Health, University College Cork, Ireland

^2^ MRC Integrative Epidemiology Unit at the University of Bristol, Bristol, UK

^3^ Population Health Sciences, Bristol Medical School Bristol, UK

**Corresponding author:** Dr Kate O’Neill, School of Public Health, 4^th^ Floor Western Gateway Building, University College Cork, Ireland

**Email**

**Supplementary Material Contents**

**Appendix 1:** Mitochondrial DNA haplogroup derivation

**Figure S1:** Flow diagram of study

**Appendix 2:** Details of measurement sources

**Appendix 3:** Details of model selection

**Table S1:** Frequency of Mitochondrial DNA Haplogroups in full sample

**Table S2:** Model details for log BMI trajectories

**Table S3:** Model details for log fat mass trajectories

**Table S4:** Model details for lean mass trajectories

**Table S5:** Model details for SBP, DBP and pulse rate trajectories

**Table S6:** Model details for HDL-c

**Table S7:** Model details for non-HDL-c trajectories

**Table S8:** Model details for log triglyceride trajectories

**Table S9**: Frequencies of haplogroups among participants with at least 1 measure of each risk factor

**Table S10:** Number of participants with cardiometabolic risk factor measures at each time point

**Table S11:** Characteristics at birth of the mothers of children included in models compared with those excluded due to missing exposure or outcome data

**Table S12:** Mean trajectories of BMI estimated from multilevel models, by haplogroup

**Table S13**: Mean trajectories of fat mass estimated from multilevel models, by haplogroup

**Table S14:** Mean trajectories of lean mass estimated from multilevel models, by haplogroup

**Table S15:** Mean trajectories of SBP, DBP and pulse rate estimated from multilevel models, by haplogroup

**Table S16:** Mean trajectories of HDL-c and non-HDL-c estimated from multilevel models**,** by haplogroup

**Table S17:** Mean trajectories of triglyceride estimated from multilevel models**,** by haplogroup

**Appendix 1:** Mitochondrial Haplogroup derivation

A total of 9,912 participants were genotyped using the Illumina HumanHap550 quad genome-wide SNP genotyping platform by Sample Logistics and Genotyping Facilities at the Wellcome Trust Sanger Institute and LabCorp (Laboratory Corporation of America) using support from 23andMe. Individuals were excluded from further analysis on the basis of having incorrect gender assignments, minimal or excessive heterozygosity (<0.320 and >0.345 for the Sanger data and <0.310 and >0.330 for the LabCorp data), disproportionate levels of individual missingness (>3%), evidence of cryptic relatedness (>10% IBD) and being of non-European ancestry (as detected by a multidimensional scaling analysis seeded with HapMap 2 individuals). EIGENSTRAT analysis revealed no additional obvious population stratification and genome-wide analyses with other phenotypes indicate a low lambda). SNPs with a minor allele frequency of <1% and call rate of <95% were removed. After QC, 8,365 unrelated individuals were available for analysis. 7,554 custom mitochondrial probes, targeting 2,824 unique mtDNA positions, were included on the Illumina HumanHap550 quad genome-wide SNP genotyping platform. All heterozygous genotype calls (i.e. heteroplasmy) were set to missing prior to quality control using PLINK (1). Genotype calls obtained from each probe were compared to the human mitochondrial database of non-pathological mitochondrial sequence variants (www.hmtdb.uniba.it:8080/hmdb/) to ensure that known allelic variants were being called. Probes were excluded in cases where genotype calls were not represented in the Cambridge Reference Sequence reference (rCRS) or one of the known allelic variants. Probes with an overall call rate of <95% were excluded prior to analysis. The genotyping concordance of the remaining probes was investigated by comparing the genotype calls in 445 replicate samples. With the exception of probe failure (i.e. missing data), a 100% genotyping concordance rate was obtained for each probe. All probes with a failure rate of >5% in the replicate sample were further excluded. In cases where multiple probes passed the above-mentioned QC criteria, the probe with the highest calling rate was used for analysis. A total of 1,062 probes passed QC for the batch that was genotyped by Laboratory Corporation of America (n=7,590), whilst 629 probes passed QC for the batch genotyped by the Sanger Institute (n=775). Haplogroup assignment was performed as described by Kloss-Brandstatter et al, and samples with a quality score of more than 90% were used for our analysis (2). Major haplogroups were defined as containing multiple haplogroups that are closely related to utilize information on less common haplogroups. After QC and removing individuals with withdrawn consent; our dataset contained 8,209 individuals with derived mitochondrial DNA haplogroups. Individuals with rare European and non-European haplogroups were excluded from our analysis (A, C, D, L, M, N, R, V; n=107). Figure S1 shows a flow diagram of the study sample.

**Figure S1: Flow diagram of study**

**Genotyping**

**Mitochondrial DNA Haplogroup determination**

**14,901 eligible participants at 1 year of age**

**Recruitment**

**Cardiometabolic risk factors**

**9,912 participants genotyped**

**Triglycerides**
**N**

**individuals**

7,172

**N measures**

16,897

**BMI**

**N individuals**

7,954

**N measures**

79,178

**Fat Mass**
**N individuals**

6,360

**N measures** 22,864

**DBP**
**N individuals**

6,913

**N measures** 34,726

**4,989 participants excluded due to loss to follow/no blood or saliva sample**

**1,703 participants excluded after quality control and withdrawn consent**

**107 participants excluded with rare European and non-European haplogroups**

**8,209 participants with derived mitochondrial DNA haplogroups**

**8,102 participants with common European haplogroups**

**Lean Mass**
**N individuals**

6,367

**N measures** 22,913

**SBP**
**N individuals**

6,912

**N measures** 34,720

**Pulse**
**N**

**individuals**

6,914

**N measures** 34,753

**Non-HDL-c**
**N individuals**

7,172

**N measures** 16,897

**HDL-c**
**N individuals**

7,176

**N measures** 16,899

BMI, body mass index; SBP, systolic blood pressure; DBP, diastolic blood pressure; HDL-c, high density lipoprotein cholesterol.

**Appendix 2:** Details of measurement sources

**Details of measurement sources**

*Details on measurement of anthropometry*

Data from age 1 onwards are included in this analysis. We did not include measures before 1 year because of the difficulty in accurately modelling BMI from birth through the whole of childhood due to its early peak followed by adiposity rebound. From 1 to 5 years, measures were available from routine child health clinics for most children and extracted from health visitor records, which form part of standard child care in the UK. Data were also available from research clinic measurements on a random 10% subsample of the cohort. All cohort members were invited to research clinics from age 7 onwards. Across all ages parent-reported measures were available.

At the clinics, crown-heel length for children aged four to 25 months was measured using a Harpenden Neonatometer and from 25 months onwards standing height was measured using a Leicester Height Measure; weight was measured using Fereday 100kg combined scale (four-month clinic), Soenhle scale or Seca scale model 724 (eight-month clinic), Seca 724 or Seca 835 (12-month clinic), Seca 835 (18 months onwards). From age 7 years, all children were invited to annual clinics, at which standing height was measured to the last complete mm using the Harpenden Stadiometer and weight was measured to the nearest 0.1kg using the Tanita Body Fat Analyser (Model TBF 305).

*Details on measurement of blood pressure*

A Dinamap 9301 Vital Signs Monitor (Morton Medical, London) was used at the 7, 9, and 11-year clinics; an Omron MI-5 was used at the 10-year clinic; a Dinamap 8100 Vital Signs Monitor (Morton Medical) was used at the 13-year clinic; and an Omron IntelliSense M6 (Omron Healthcare, Kyoto, Japan) was used at the 15- and 18-year clinics.

*Details on measurement of blood-based biomarkers*

Plasma lipid assays (triglycerides and high-density lipoprotein cholesterol (HDL-c)) were performed by modification of the standard Lipid Research Clinics Protocol using enzymatic reagents for lipid determination. All assay coefficients of variation were <5%.

**Appendix 3:** Details of model selection

Two approaches, fractional polynomials and linear splines were used in the modelling of trajectories as described previously (3–6).

Fractional polynomials were used for body mass index (BMI), due to the complex pattern of change in BMI during childhood and adolescence. Fractional polynomials involve raising age to many combinations of powers, resulting in a wide range of possible curves and offering more flexibility than standard polynomial approaches.

Linear splines were used in the modelling of all other outcomes in this paper as too few measurement occasions were available to permit modelling using fractional polynomials. Models were derived by initially examining observed data for each risk factor. We also plotted mean values for each risk factor on each measurement occasion to assist on decisions regarding knot points. We compared observed and predicted measurements for a selection of suitable models for each risk factor. We examined rates of change between time periods in order to examine whether changes between periods were similar or different. In cases where rates of change between two spline periods appeared identical, the fit of models with a reduced number of splines was explored.

Specific details for each model are described here:

**BMI** has been modelled previously using fractional polynomials and is described elsewhere. Briefly, BMI was log transformed due to skewness of the data and fractional polynomials were used where age was raised to various combinations of powers (each of the following single powers, plus each combination of two powers: 0.5, 1, 2, 3, -0.5, -1, -2, natural log), from which we selected the best fitting curve (the one with the lowest likelihood value). The resulting curve contained three age terms including log age, log age* age and log age *age^^2.^ To account for the likely reduced accuracy of parent-reported measurements, a binary indicator of measurement source (research clinic or health records versus parent-report) was included as a fixed effect. The variance of measurement occasion-level residuals (the differences between observed and predicted measurements) was allowed to vary with age for log BMI. The model took the form of: log BMI_ij_ = (β_0_+u_0j_+e_0ij_) + (β_1_+u_1j_)(ln(age)_ij_) + (β_2_+u_2j_)(age*ln(age)_ij_) + (β_3_+u_3j_)(age^2^*ln(age)_ij_) + (β_8_+e_1ij_)(measurement_source_ij_) + e_ij_(age_months_ij_) where for person j at measurement occasion i; β’s represent fixed effect coefficients, u_0j_ to u_3j_ indicate person-specific random effects for the intercept and linear, quadratic and cubic age terms respectively, and e_1_ represents the occasion-specific residuals or measurement error which was allowed to vary with age and according to measurement source.

**Fat mass** and **lean mass** were measured on five occasions between 9 and 18 years. Fat mass was log transformed due to skewness of the data. Knots were placed at 13 and 15 years resulting in three periods of change; from 9-13, 13-15, and 15-18. Both models were adjusted for a time varying height co-variate, which was included as a fixed effect and is described elsewhere in detail. The models took the form of: log fat mass_ij_ or lean mass_ij_ = β_0_ + u_0j_ + (β_1_+ u_1j_ )s_ij1_ + (β_2_+ u_2j_ )s_ij2_ + (β_3_ + u_3j_)s_ij3_ + β_4_ (age-adjusted height covariate)_ij_ + e_ij_ where for person j at measurement occasion i; β_0_ represents the fixed effect coefficient for the average intercept, β_1_ to β_3_ represent fixed effect coefficients for the average linear slopes of each linear spline, s_ij_ represents the specific spline period, β_4_ represents the fixed effect coefficient for the average difference in measurements between individuals of different heights, u_0j_ to u_3j_ indicate person-specific random effects for the intercept and slopes respectively, and e_ij_ represents the occasion-specific residuals or measurement error which was allowed to vary with age.

**SBP, DBP** and **pulse rate** were measured at 7 time points from 7 to 18 years. The models for SBP and DBP is described elsewhere in detail (3,5). The knots for all models were placed at 12 and 16 resulting in three periods of change; from 7-12, 12-16 and 16-18. All models included a fixed effect to adjust for the use of the use of an Omron MI-5 machine to measure SBP in 10-year clinic which differed from all other clinics and a binary time indicator as a level one random effect of age less than or greater than 10 years to account for changing measurement error with age. The models took the form of: SBP_ij_ or DBP_ij_ or pulse_ij_ = β_0_ + u_0j_ + (β_1_+ u_1j_ )s_ij1_ + (β_2_+ u_2j_ )s_ij2_ + (β_3_ + u_3j_ )s_ij3_ + β_4_ (machine)_ij_ + e_ij_(age_binary_ij_) where for person j at measurement occasion i; β_0_ represents the fixed effect coefficient for the average intercept, β_1_ to β_3_ represent fixed effect coefficients for the average linear slopes of each linear spline, s_ij_ represents the specific spline period, β_4_ represents the fixed effect coefficient for the average difference in measurements between the machine used at the 10 year clinic compared to the machine used at other clinics, u_0j_ to u_3j_ indicate person-specific random effects for the intercept and slopes respectively, and e_ij_ represents the occasion-specific residuals or measurement error which was allowed to vary with age.

**HDL-c** and **Triglycerides** were measured 5 times from birth to 18 years. Triglyceride was log transformed due to the skewness of the data. **Non-HDL-c** was derived by subtracting HDL-c from total cholesterol. Knots for triglyceride and non-HDL-c were placed at 9 and 15 years resulting in 2 periods of change; from birth to 9 years and 9-18. Knots were placed at age 7 and 15 years for HDL-c resulting in two periods of change; from birth to 7 years and 7-18 years. The models took the form of: log triglycerides_ij_ or HDL-c_ij_ or non-HDL-c_ij_ = β_0_ + u_0j_ + (β_1_+ u_1j_ )s_ij1_ + (β_2_+ u_2j_ )s_ij2_ + e_ij_ where for person j at measurement occasion i; β_0_ represents the fixed effect coefficient for the average intercept, β_1_ and β_2_ represent fixed effect coefficients for the average linear slopes of each linear spline, s_ij_ represents the specific spline period, u_0j_ to u_3j_ indicate person-specific random effects for the intercept and slopes respectively, and e_ij_ represents the occasion-specific residuals or measurement error.

Observed and predicted measurements for each model are shown in Tables S2-S8.

| **Haplogroup** | **N = 8,209** | **%** |
| --- | --- | --- |
| **H** | 3,649 | 44.45 |
| **U** | 1,083 | 13.19 |
| **T** | 830 | 10.11 |
| **J** | 883 | 10.76 |
| **K** | 704 | 8.58 |
| **V** | 456 | 5.55 |
| **W** | 147 | 1.79 |
| **I** | 231 | 2.81 |
| **X** | 119 | 1.45 |
| **M** | 29 | 0.35 |
| **L** | 28 | 0.34 |
| **N** | 18 | 0.22 |
| **R** | 14 | 0.17 |
| **A** | 8 | 0.10 |
| **D** | 7 | 0.09 |
| **C** | 3 | 0.04 |

**Table S1:** Frequency of mitochondrial DNA haplogroups in full sample **Table S2:** Model details for log BMI trajectories

|  | Number of contributing individuals | | Assessment of model fit | | | |
| --- | --- | --- | --- | --- | --- | --- |
|  | Total observations | Individuals with 1 measure | Mean observed  BMI, ln(kg/m^2^) (SD) ^a^ | Mean predicted  BMI, ln(kg/m^2^) (SD) ^a^ | Mean difference (observed – predicted), ln(kg/m^2^) ^a^ | 95% level of agreement between observed and predicted, ln(kg/m^2^) ^a^ |
| Female |  |  |  |  |  |  |
| Overall | 39778 | 3886 |  |  |  |  |
| 1-3 years | 6307 | 3034 | 2.81 (0.09) | 2.81 (0.07) | 0.004 (0.07) | -0.09 to 0.10 |
| 3-7 years | 8328 | 3320 | 2.77 (0.11) | 2.77 (0.08) | -0.001 (0.08) | -0.14 to 0.14 |
| 7-9 years | 5272 | 3055 | 2.81 (0.13) | 2.82 (0.12) | -0.01 (0.12) | -0.08 to 0.06 |
| 9-11 years | 6078 | 3065 | 2.88 (0.16) | 2.88 (0.14) | -0.001 (0.14) | -0.08 to 0.08 |
| 11-13 years | 5018 | 2876 | 2.95 (0.17) | 2.94 (0.15) | 0.01 (0.15) | -0.07 to 0.10 |
| 13-15 years | 4610 | 2754 | 3.00 (0.16) | 2.99 (0.15) | 0.01 (0.15) | -0.11 to 0.12 |
| 15-18 years | 4165 | 2478 | 3.08 (0.15) | 3.10 (0.15) | -0.01 (0.15) | -0.10 to 0.08 |
| Male |  |  |  |  |  |  |
| Overall | 39400 | 4068 |  |  |  |  |
| 1-3 years | 6653 | 3207 | 2.84 (0.09) | 2.83 (0.07) | 0.01 (0.07) | -0.09 to 0.10 |
| 3-7 years | 8811 | 3525 | 2.78 (0.10) | 2.78 (0.07) | -0.002 (0.07) | -0.13 to 0.12 |
| 7-9 years | 5297 | 3104 | 2.79 (0.12) | 2.80 (0.11) | -0.01 (0.11) | -0.08 to 0.06 |
| 9-11 years | 5978 | 3007 | 2.86 (0.15) | 2.86 (0.14) | 0.002 (0.14) | -0.07 to 0.07 |
| 11-13 years | 4666 | 2762 | 2.93 (0.16) | 2.92 (0.15) | 0.01 (0.15) | -0.06 to 0.09 |
| 13-15 years | 4441 | 2676 | 2.97 (0.16) | 2.97 (0.15) | 0.001 (0.15) | -0.10 to 0.10 |
| 15-18 years | 3554 | 2181 | 3.06 (0.15) | 3.07 (0.15) | -0.01 (0.15) | -0.09 to 0.07 |

SD, standard deviation; ln(kg/m^2^), natural log of kilograms per metre squared

^a^ BMI is presented in the natural log and values represent the mean predicted natural log of BMI at each age shown.

**Table S3:** Model details for log fat mass trajectories

|  | **Number of contributing individuals** | | **Assessment of model fit** | | | |
| --- | --- | --- | --- | --- | --- | --- |
|  | Total observations | Individuals with 1 measure | Mean observed, ln(kg) (SD)^a^ | Mean predicted, ln(kg) (SD)^aa^ | Mean difference (observed – predicted), ln(kg)^a^ | 95% level of agreement between observed and predicted, ln(kg)^a^ |
| **Female** |  |  |  |  |  |  |
| Overall | 11909 | 3216 |  |  |  |  |
| 9 years | 2803 | 2803 | 2.14 (0.50) | 2.14 (0.48) | -0.003 | -0.15 to 0.14 |
| 9-13 years | 5496 | 3083 | 2.28 (0.52) | 2.28 (0.49) | 0.002 | -0.17 to 0.17 |
| 13-15 years | 2394 | 2373 | 2.69 (0.44) | 2.70 (0.42) | -0.01 | -0.21 to 0.20 |
| 15-18 years | 4019 | 2421 | 2.91 (0.41) | 2.91 (0.38) | 0.002 | -0.16 to 0.16 |
| **Male** |  |  |  |  |  |  |
| Overall | 10955 | 3144 |  |  |  |  |
| 9 years | 2705 | 2705 | 1.81 (0.59) | 1.83 (0.55) | -0.02 | -0.23 to 0.20 |
| 9-13 years | 5274 | 3009 | 1.98 (0.62) | 1.97 (0.57) | 0.01 | -0.23 to 0.26 |
| 13-15 years | 2238 | 2228 | 2.20 (0.62) | 2.25 (0.57) | -0.05 | -0.36 to 0.27 |
| 15-18 years | 3443 | 2125 | 2.31 (0.64) | 2.30 (0.60) | 0.01 | -0.23 to 0.25 |

SD, standard deviation; ln(kg), natural log of kilograms

^a^Fat mass is presented in the natural log and values represent the mean predicted natural log of fat mass.

**Table S4:** Model details for lean mass trajectories

|  | **Number of contributing individuals** | | **Assessment of model fit** | | | | |
| --- | --- | --- | --- | --- | --- | --- | --- |
|  | **Total observations** | **Individuals with 1 measure** | **Mean observed, kg (SD)** | **Mean predicted, kg (SD)** | | **Mean difference (observed – predicted), kg** | **95% level of agreement between observed and predicted, kg** |
| **Female** |  |  |  |  |  | |  |
| Overall | 11940 | 3220 |  |  |  | |  |
| 9 years | 2807 | 2807 | 23.61 (3.11) | 23.55 (2.94) | 0.06 | | -2.28 to 2.40 |
| 9-13 years | 5506 | 3087 | 26.40 (4.74) | 26.46 (4.51) | -0.07 | | -2.38 to 2.25 |
| 13-15 years | 2402 | 2381 | 35.27 (4.00) | 35.01 (3.85) | 0.26 | | -2.06 to 2.58 |
| 15-18 years | 4032 | 2428 | 37.55 (4.06) | 37.62 (3.75) | -0.06 | | -2.05 to 1.93 |
| **Male** |  |  |  |  |  | |  |
| Overall | 10973 | 3147 |  |  |  | |  |
| 9 years | 2711 | 2711 | 25.54 (2.91) | 25.38 (2.39) | 0.16 | | -3.56 to 3.88 |
| 9-13 years | 5283 | 3013 | 27.80 (4.26) | 27.95 (4.21) | -0.14 | | -3.49 to 3.20 |
| 13-15 years | 2240 | 2230 | 40.95 (7.12) | 40.39 (6.08) | 0.55 | | -3.36 to 4.47 |
| 15-18 years | 3450 | 2129 | 52.24 (7.06) | 52.38 (6.69) | -0.14 | | -2.86 to 2.59 |

SD, standard deviation; kg, kilograms

**Table S5:** Model details for SBP, DBP and pulse rate trajectories

|  | Number of contributing individuals | | Assessment of model fit | | | | |
| --- | --- | --- | --- | --- | --- | --- | --- |
|  | Total observations | Individuals with 1 measure | Mean observed SBP, DBP or pulse rate (SD) ^a^ | Mean predicted SBP, DBP or pulse rate (SD) ^a^ | Mean difference (observed–predicted) ^a^ | | 95% level of agreement between observed and predicted ^a^ |
| SBP |  |  |  |  | |  |  |
| Females |  |  |  |  | |  |  |
| Overall | 17863 | 3453 |  |  | |  |  |
| 7 years | 2926 | 2926 | 98.95 (9.25) | 98.79 (5.57) | | 0.16 | -10.59 to 10.92 |
| 7-12 years | 11035 | 3373 | 102.88 (9.75) | 103.02 (6.34) | | -0.13 | -11.98 to 11.72 |
| 12-16 years | 4776 | 2768 | 114.66 (11.04) | 114.20 (7.79) | | 0.46 | -11.31 to 12.23 |
| 16-18 years | 2052 | 1986 | 112.54 (8.48) | 112.87 (5.44) | | -0.33 | -12.72 to 12.06 |
| Males |  |  |  |  | |  |  |
| Overall | 16857 | 3459 |  |  | |  |  |
| 7 years | 2989 | 2989 | 98.68 (9.04) | 98.61 (5.34) | | 0.08 | -10.43 to 10.58 |
| 7-12 years | 10801 | 3392 | 102.29 (9.34) | 102.41 (6.12) | | -0.12 | -11.47 to 11.23 |
| 12-16 years | 4450 | 2626 | 116.94 (12.41) | 116.53 (9.68) | | 0.40 | -11.46 to 12.27 |
| 16-18 years | 1606 | 1560 | 122.38 (9.48) | 122.67 (5.93) | | -0.29 | -12.07 to 11.48 |
| DBP  Females |  |  |  |  | |  |  |
| Overall | 17869 | 3454 |  |  | |  |  |
| 7 years | 2926 | 2926 | 56.69 (6.53) | 56.91 (3.37) | | -0.22 | -9.00 to 8.55 |
| 7-12 years | 11045 | 3373 | 58.45 (7.01) | 58.01 (3.71) | | 0.44 | -9.71 to 10.59 |
| 12-16 years | 4776 | 2768 | 60.97 (9.25) | 61.86 (4.70) | | -0.89 | -12.56 to 10.77 |
| 16-18 years  Males | 2048 | 1981 | 64.65 (5.87) | 64.94 (3.47) | | -0.29 | -13.13 to 12.55 |
| Overall | 16857 | 3459 |  |  | |  |  |
| 7 years | 2988 | 2988 | 55.94 (6.63) | 56.19 (3.43) | | -0.24 | -8.70 to 8.21 |
| 7-12 years | 10802 | 3392 | 57.60 (6.86) | 57.19 (3.56) | | 0.41 | -9.42 to 10.23 |
| 12-16 years | 4451 | 2626 | 61.35 (10.12) | 62.23 (5.67) | | -0.88 | -12.49 to 10.73 |
| 16-18 years | 1604 | 1559 | 63.44 (6.17) | 63.82 (3.72) | | -0.38 | -12.81 to 12.06 |
| Pulse  Females |  |  |  |  | |  |  |
| Overall | 17888 | 3455 |  |  | |  |  |
| 7 years | 2920 | 2927 | 84.31 (10.75) | 84.83 (5.74) | | -0.52 | -14.37 to 13.34 |
| 7-12 years | 10828 | 3373 | 79.69 (11.28) | 79.33 (7.19) | | 0.27 | -14.03 to 14.57 |
| 12-16 years | 4765 | 2769 | 75.80 (11.04) | 76.29 (6.30) | | -0.50 | -14.74 to 13.74 |
| 16-18 years | 2052 | 1993 | 67.58 (9.98) | 67.85 (5.90) | | -0.28 | -14.93 to 14.37 |
| Pulse  Males |  |  |  |  | |  |  |
| Overall | 16865 | 3459 |  |  | |  |  |
| 7 years | 2990 | 2991 | 81.68 (10.57) | 81.94 (6.03) | | -0.26 | -11.79 to 11.28 |
| 7-12 years | 10406 | 3392 | 76.20 (11.31) | 76.04 (7.63) | | 0.003 | -12.93 to 12.94 |
| 12-16 years | 4443 | 2626 | 72.15 (11.13) | 72.07 (6.67) | | 0.07 | -13.32 to 13.47 |
| 16-18 years | 1606 | 1561 | 63.07 (9.53) | 63.30 (5.38) | | -0.22 | -14.55 to 14.10 |

DBP, diastolic blood pressure; SBP, systolic blood pressure; SD, standard deviation

^a^units are presented in mmHg for SBP and DBP and bpm for pulse rate

**Table S6:** Model details for HDL-c trajectories

|  | **Number of contributing individuals** | | **Assessment of model fit** | | | |
| --- | --- | --- | --- | --- | --- | --- |
|  | Total observations | Individuals with 1 measure | Mean observed, mmol/l (SD) | Mean predicted, mmol/l (SD) | Mean difference (observed – predicted), mmol/l | 95% level of agreement between observed and predicted, mmol/l |
| **Females** |  |  |  |  |  |  |
| Overall | 8463 | 3533 |  |  |  |  |
| Birth | 1474 | 1474 | 0.84 (3.98) | 0.84 (3.97) | 0.00000002 | -0.02 to 0.02 |
| 0-7 years | 3488 | 2799 | 1.22 (2.62) | 1.20 (2.60) | 0.02 | -0.18 to 0.22 |
| 7-18 years | 6988 | 3099 | 1.40 (0.31) | 1.40 (0.22) | 0.0000 | -0.28 to 0.28 |
| **Male** |  |  |  |  |  |  |
| Overall | 8436 | 3643 |  |  |  |  |
| Birth | 1515 | 1515 | 0.69 (2.98) | 0.69 (2.97) | -0.000002 | -0.02 to 0.02 |
| 0-7 years | 3640 | 2969 | 1.19 (1.98) | 1.18 (1.97) | 0.005 | -0.17 to 0.18 |
| 7-18 years | 6920 | 3133 | 1.38 (0.32) | 1.38 (0.26) | -0.00002 | -0.25 to 0.25 |

HDL-c, high density lipoprotein cholesterol; SD, standard deviation; mmol/l, millimole per litre

**Table S7:** Model details for non-HDL-c trajectories

|  | **Number of contributing individuals** | | **Assessment of model fit** | | | |
| --- | --- | --- | --- | --- | --- | --- |
|  | Total observations | Individuals with 1 measure | Mean observed, mmol/l (SD) | Mean predicted, mmol/l (SD) | Mean difference (observed – predicted), mmol/l | 95% level of agreement between observed and predicted, mmol/l |
| **Females** |  |  |  |  |  |  |
| Overall | 8458 | 3529 |  |  |  |  |
| Birth | 1474 | 1474 | 1.07 (4.12) | 1.09 (4.00) | -0.02 | -0.26 to 0.21 |
| 0-7 years | 3575 | 2872 | 2.19 (2.85) | 2.15 (2.74) | 0.05 | -0.62 to 0.71 |
| 7-18 years | 4883 | 2645 | 2.74 (0.68) | 2.78 (0.51) | -0.03 | -0.81 to 0.74 |
| **Males** |  |  |  |  |  |  |
| Overall | 8439 | 3643 |  |  |  |  |
| Birth | 1515 | 1515 | 1.07 (3.11) | 1.08 (3.06) | -0.02 | -0.15 to 0.12 |
| 0-7 years | 3730 | 3040 | 2.08 (2.20) | 2.05 (2.13) | 0.03 | -0.37 to 0.44 |
| 7-18 years | 4709 | 2587 | 2.55 (0.64) | 2.58 (0.51) | -0.03 | -0.56 to 0.51 |

Non-HDL-c, non-high density lipoprotein cholesterol; SD, standard deviation; mmol/l, millimole per litre

**Table S8:** Model details for log triglyceride trajectories

|  | **Number of contributing individuals** | | **Assessment of model fit** | | | |
| --- | --- | --- | --- | --- | --- | --- |
|  | Total observations | Individuals with 1 measure | Mean observed, ln(trig), (SD)^a^ | Mean predicted, ln(trig), (SD)^a^ | Mean difference (observed – predicted), ln(trig)^a^ | 95% level of agreement between observed and predicted, ln(trig)^a^ |
| **Females** |  |  |  |  |  |  |
| Overall | 8458 | 3529 |  |  |  |  |
| Birth | 1474 | 1474 | -0.68 (0.45) | -0.67 (0.22) | -0.005 | -0.68 (0.45) |
| 0-9 years | 3575 | 2872 | -0.29 (0.54) | -0.30 (0.38) | 0.01 | -0.29 (0.54) |
| 9-18 years | 4883 | 2645 | -0.11 (0.41) | -0.11 (0.23) | -0.01 | -0.11 (0.41) |
| **Males** |  |  |  |  |  |  |
| Overall | 8439 | 3643 |  |  |  |  |
| Birth | 1515 | 1515 | -0.68 (0.45) | -0.68 (0.19) | -0.002 | -0.68 (0.45) |
| 0-9 years | 3730 | 3040 | -0.33 (0.53) | -0.33 (0.34) | 0.003 | -0.33 (0.53) |
| 9-18 years | 4709 | 2587 | -0.15 (0.42) | -0.15 (0.22) | -0.003 | -0.15 (0.42) |

ln(trig), natural log of triglyceride; SD, standard deviation

^a^Triglyceride is presented in the natural log and values represent the mean predicted natural log of triglyceride at each age shown.

**Table S9:** Frequencies of haplogroups among participants with at least 1 measure of each risk factor

|  | BMI  n (%) | Fat mass  n (%) | Lean mass  n (%) | SBP  n (%) | DBP  n (%) | Pulse  n (%) | HDL-c  n (%) | Non-HDL-c  n (%) | Triglycerides  n (%) |
| --- | --- | --- | --- | --- | --- | --- | --- | --- | --- |
| **Females** |  |  |  |  |  |  |  |  |  |
| **Total** | 3886 | 3216 | 3220 | 3453 | 3454 | 3455 | 3533 | 3529 | 3529 |
| **H** | 1759 (45.3) | 1449 (45.1) | 1452 (45.1) | 1570 (45.5) | 1570 (45.5) | 1571 (45.5) | 1600 (45.3) | 1598 (45.3) | 1597 (45.3) |
| **U** | 515 (13.3) | 434 (13.5) | 434 (13.5) | 457 (13.2) | 457 (13.2) | 457 (13.2) | 473 (13.4) | 472 (13.4) | 472 (13.4) |
| **T** | 401 (10.3) | 341 (10.6) | 341 (10.6) | 357 (10.3) | 357 (10.3) | 357 (10.3) | 370 (10.5) | 370 (10.5) | 368 (10.4) |
| **J** | 420 (10.8) | 353 (11.0) | 354 (11.0) | 377 (10.9) | 377 (10.9) | 377 (10.9) | 389 (11.0) | 389 (11.0) | 389 (11.0) |
| **K** | 300 (7.7) | 247 (7.7) | 247 (7.7) | 272 (7.9) | 272 (7.9) | 272 (7.9) | 277 (7.8) | 277 (7.8) | 278 (7.9) |
| **V** | 243 (6.3) | 193 (6.0) | 193 (6.0) | 209 (6.1) | 209 (6.1) | 209 (6.0) | 214 (6.1) | 214 (6.1) | 214 (6.1) |
| **W** | 67 (1.7) | 52 (1.6) | 52 (1.6) | 55 (1.6) | 55 (1.6) | 55 (1.6) | 58 (1.6) | 58 (1.6) | 58 (1.6) |
| **I** | 125 (3.2) | 99 (3.1) | 99 (3.1) | 105 (3.0) | 105 (3.0) | 105 (3.0) | 102 (2.9) | 102 (2.9) | 103 (2.9) |
| **X** | 56 (1.4) | 48 (1.5) | 48 (1.5) | 51 (1.5) | 52 (1.5) | 52 (1.5) | 50 (1.4) | 49 (1.4) | 50 (1.4) |
| **Males** |  |  |  |  |  |  |  |  |  |
| **Total** | 4068 | 3144 | 3147 | 3459 | 3459 | 3459 | 3643 | 3643 | 3643 |
| **H** | 1818 (44.7) | 1389 (44.2) | 1391 (44.2) | 1538 (44.5) | 1538 (44.5) | 1538 (44.5) | 1630 (44.7) | 1630 (44.7) | 1629 (44.7) |
| **U** | 548 (13.5) | 421 (13.4) | 421 (13.4) | 456 (13.2) | 456 (13.2) | 456 (13.2) | 484 (13.3) | 484 (13.3) | 487 (13.4) |
| **J** | 415 (10.2) | 330 (10.5) | 330 (10.5) | 362 (10.5) | 362 (10.5) | 362 (10.5) | 371 (10.2) | 371 (10.2) | 371 (10.2) |
| **T** | 448 (11.0) | 350 (11.1) | 351 (11.2) | 384 (11.1) | 384 (11.1) | 384 (11.1) | 407 (11.2) | 407 (11.2) | 407 (11.2) |
| **K** | 390 (9.6) | 299 (9.5) | 299 (9.5) | 325 (9.4) | 325 (9.4) | 325 (9.4) | 344 (9.4) | 344 (9.4) | 342 (9.4) |
| **V** | 205 (5.0) | 163 (5.2) | 163 (5.2) | 178 (5.1) | 178 (5.1) | 178 (5.1) | 185 (5.1) | 185 (5.1) | 185 (5.1) |
| **W** | 80 (2.0) | 61 (1.9) | 61 (1.9) | 71 (2.1) | 71 (2.1) | 71 (2.1) | 72 (2.0) | 72 (2.0) | 72 (2.0) |
| **I** | 101 (2.5) | 81 (2.6) | 81 (2.6) | 92 (2.7) | 92 (2.7) | 92 (2.7) | 93 (2.6) | 93 (2.6) | 93 (2.6) |
| **X** | 63 (1.5) | 50 (1.6) | 50 (1.6) | 53 (1.5) | 53 (1.5) | 53 (1.5) | 57 (1.6) | 57 (1.6) | 57 (1.6) |

**Table S10:** Number of participants with cardiometabolic risk factor measures at each time point

|  | | Birth | | Age 1 | | Age 7 | | Age 9 | | Age 10 | | Age 11 | | | Age 12 | | Age 13 | | Age 15 | | Age 18 |
| --- | --- | --- | --- | --- | --- | --- | --- | --- | --- | --- | --- | --- | --- | --- | --- | --- | --- | --- | --- | --- | --- |
| BMI ^a^ | | |  | x | x | | x | | x | | x | | x | | | x | | x | | x | |
| Fat mass | | |  |  |  | | 5,508 | |  | | 5,256 | |  | | | 4,605 | | 3,942 | | 3,553 | |
| Lean mass | | |  |  |  | | 5,518 | |  | | 5,265 | |  | | | 4,615 | | 3,955 | | 3,560 | |
| SBP | | |  |  | 5,915 | | 5,714 | | 5,356 | | 5,279 | | 4,996 | | |  | | 4,029 | | 3,431 | |
| DBP | | |  |  | 5,914 | | 5,722 | | 5,356 | | 5,284 | | 4,996 | | |  | | 4,030 | | 3,424 | |
| Pulse rate | | |  |  | 5,918 | | 5,719 | | 5,362 | | 5,288 | | 4,996 | | |  | | 4,031 | | 3,439 | |
| HDL | | | 2,989 |  | 4,387 | | 4,184 | |  | |  | |  | | |  | | 2,829 | | 2,510 | |
| Non-HDL | | | 2,989 |  | 4,387 | | 4,182 | |  | |  | |  | | |  | | 2,829 | | 2,510 | |
| Triglycerides | | | 3,015 |  | 4,366 | | 4,161 | |  | |  | |  | | |  | | 2,817 | | 2,494 | |

BMI, body mass index; HDL, high density lipoprotein cholesterol; DBP, diastolic blood pressure; SBP, systolic blood pressure.

^a^ Measures available at each of these approximate ages and at several ages in between but exact timing and number of BMI measures not shown as measures were available from questionnaires, routine child health records and research clinics at different mean ages from 1 to 18 years.

**Table S11:** Characteristics at birth of the mothers of children included in models compared with those excluded due to missing exposure or outcome data

|  | **Included**  **N=6,794-7,387^a^** | **Excluded**  **N=4,673-6,037^a^** |
| --- | --- | --- |
|  | **n (%)** | **n (%)** |
| **Marital Status** |  |  |
| Never | 1119(15.2) | 1442(23.9) |
| Widowed | 11(0.1) | 7(0.1) |
| Divorced | 286(3.9) | 284(4.7) |
| Separated | 100(1.4) | 116(1.9) |
| 1st marriage | 5336(72.5) | 3814(63.2) |
| 2 or 3 marriage | 505(6.9) | 374(6.2) |
| **Household Social Class** † |  |  |
| Professional | 1005(14.8) | 516(11.0) |
| Managerial/technical | 3003(44.2) | 1797(38.5) |
| Non manual | 1691(24.9) | 1230(26.3) |
| Manual | 764(11.2) | 775(16.6) |
| Part skilled and unskilled | 331(4.9) | 355(7.6) |
| **Maternal Education** |  |  |
| Less than O level | 1784(24.9) | 1932(37.2) |
| O level | 2502(34.9) | 1780(34.3) |
| A level | 1791(25.0) | 990(19.1) |
| Degree or above | 1099(15.3) | 492(9.5) |
| **Mother’s Partner’s Education** |  |  |
| Less than O level | 2075(29.8) | 2039(41.4) |
| O level | 1517(21.8) | 1009(20.5) |
| A level | 1908(27.4) | 1185(24.0) |
| Degree or above | 1467(21.1) | 695(14.1) |
| **Maternal Smoking during Pregnancy** |  |  |
| No | 5822(78.8) | 4056(69.8) |
| Yes | 1565(21.2) | 1755(30.2) |

^a^ Denominators for excluded participants in this table vary due to different rates of missing data for characteristics shown.

† Household social class was measured as the highest of the mother’s or her partner’s occupational social class using data on job title and details of occupation. Social class was derived using the standard occupational classification (SOC) codes developed by the United Kingdom Office of Population Census and Surveys and classified as I professional, II managerial and technical, IIINM non-manual, IIIM manual, and IV&V part skilled occupations and unskilled occupations.

**Table S12:** Mean trajectories of BMI and mean differences by haplogroup, estimated from multilevel models

|  | **Mean trajectory (95% CI) in haplogroup H (reference)^a^** | **Mean difference in trajectory (95% CI) comparing with haplogroup H^b^** | | | | | | | |
| --- | --- | --- | --- | --- | --- | --- | --- | --- | --- |
|  |  | **Haplogroup U** | **Haplogroup T** | **Haplogroup J** | **Haplogroup K** | **Haplogroup V** | **Haplogroup W** | **Haplogroup I** | **Haplogroup X** |
| **Females** |  |  |  |  |  |  |  |  |  |
| Age 1yr | 2.92 (2.91,2.93) | 0.02 (-1.99,2.04) | -1.24 (-3.39,0.91) | -1.83 (-4.06,0.39) | -2.07 (-4.48,0.34) | -4.02 (-6.66,-1.38) | 2.85 (-2.89,8.60) | 0.43 (-3.17,4.03) | -2.04 (-6.93,2.84) |
| Age 3yr | 2.75 (2.74,2.75) | -0.12 (-1.03,0.80) | 0.29 (-0.72,1.29) | -0.36 (-1.36,0.64) | -0.30 (-1.44,0.84) | -0.58 (-1.83,0.68) | -3.06 (-5.38,-0.74) | 0.93 (-0.78,2.65) | -2.33 (-4.71,0.06) |
| Age 7yr | 2.80 (2.79,2.80) | -0.06 (-1.23,1.10) | 0.49 (-0.80,1.78) | -0.18 (-1.44,1.08) | -0.77 (-2.21,0.68) | -2.10 (-3.68,-0.53) | -2.64 (-5.56,0.27) | 1.14 (-1.04,3.33) | -3.34 (-6.34,-0.34) |
| Age 9yr | 2.85 (2.84,2.86) | -0.03 (-1.43,1.37) | 0.42 (-1.13,1.98) | -0.19 (-1.70,1.33) | -1.07 (-2.80,0.66) | -2.82 (-4.70,-0.93) | -2.03 (-5.56,1.50) | 1.25 (-1.39,3.88) | -3.55 (-7.16,0.05) |
| Age 11yr | 2.91 (2.90,2.92) | -0.01 (-1.59,1.58) | 0.31 (-1.44,2.06) | -0.19 (-1.90,1.52) | -1.30 (-3.25,0.65) | -3.22 (-5.34,-1.10) | -1.46 (-5.47,2.55) | 1.37 (-1.61,4.34) | -3.50 (-7.57,0.56) |
| Age 13yr | 2.98 (2.97,2.99) | 0.01 (-1.65,1.68) | 0.16 (-1.68,2.00) | -0.17 (-1.97,1.63) | -1.43 (-3.48,0.62) | -3.24 (-5.47,-1.01) | -1.02 (-5.26,3.22) | 1.51 (-1.63,4.66) | -3.17 (-7.48,1.13) |
| Age 15yr | 3.05 (3.04,3.06) | 0.02 (-1.67,1.72) | -0.02 (-1.88,1.84) | -0.12 (-1.96,1.72) | -1.44 (-3.52,0.64) | -2.82 (-5.10,-0.54) | -0.75 (-5.09,3.59) | 1.68 (-1.53,4.90) | -2.54 (-6.97,1.89) |
| Age 18yr | 3.17 (3.16,3.18) | 0.02 (-1.92,1.95) | -0.33 (-2.42,1.76) | 0.02 (-2.09,2.14) | -1.19 (-3.57,1.18) | -1.29 (-3.94,1.37) | -0.77 (-5.81,4.28) | 2.00 (-1.70,5.69) | -0.97 (-6.14,4.19) |
| **Males** |  |  |  |  |  |  |  |  |  |
| Age 1yr | 2.91 (2.91,2.92) | 0.53 (-1.19,2.25) | -1.40 (-3.24,0.43) | 0.99 (-0.92,2.91) | -0.51 (-2.45,1.43) | 0.97 (-1.72,3.66) | -0.08 (-3.96,3.80) | 2.49 (-1.13,6.12) | 0.10 (-4.09,4.29) |
| Age 3yr | 2.77 (2.76,2.77) | -0.22 (-1.02,0.59) | 0.21 (-0.69,1.11) | -0.37 (-1.25,0.50) | 0.21 (-0.72,1.14) | -0.85 (-2.06,0.36) | 0.98 (-0.91,2.86) | 1.29 (-0.41,2.99) | 1.18 (-0.93,3.28) |
| Age 7yr | 2.79 (2.78,2.79) | 0.57 (-0.48,1.62) | -0.41 (-1.56,0.74) | -0.45 (-1.57,0.67) | 1.02 (-0.18,2.22) | -1.41 (-2.96,0.15) | 1.68 (-0.76,4.13) | 2.40 (0.22,4.59) | 0.36 (-2.38,3.10) |
| Age 9yr | 2.83 (2.82,2.83) | 0.84 (-0.46,2.14) | -0.68 (-2.09,0.73) | -0.38 (-1.76,1.00) | 1.26 (-0.23,2.75) | -1.52 (-3.43,0.40) | 1.96 (-1.07,4.98) | 3.06 (0.36,5.75) | 0.07 (-3.30,3.45) |
| Age 11yr | 2.88 (2.87,2.89) | 0.92 (-0.58,2.42) | -0.79 (-2.42,0.83) | -0.29 (-1.89,1.30) | 1.38 (-0.34,3.09) | -1.57 (-3.78,0.63) | 2.19 (-1.31,5.69) | 3.67 (0.54,6.79) | -0.02 (-3.90,3.87) |
| Age 13yr | 2.95 (2.94,2.95) | 0.79 (-0.83,2.41) | -0.72 (-2.48,1.05) | -0.21 (-1.94,1.52) | 1.36 (-0.50,3.22) | -1.59 (-3.98,0.79) | 2.40 (-1.41,6.22) | 4.20 (0.80,7.59) | 0.12 (-4.08,4.32) |
| Age 15yr | 3.02 (3.01,3.03) | 0.43 (-1.28,2.13) | -0.42 (-2.28,1.45) | -0.15 (-1.97,1.68) | 1.21 (-0.75,3.16) | -1.58 (-4.09,0.93) | 2.58 (-1.49,6.66) | 4.63 (1.05,8.21) | 0.51 (-3.91,4.93) |
| Age 18yr | 3.16 (3.15,3.17) | -0.59 (-2.60,1.42) | 0.47 (-1.77,2.72) | -0.08 (-2.27,2.11) | 0.70 (-1.61,3.01) | -1.51 (-4.48,1.46) | 2.80 (-2.17,7.77) | 5.06 (0.85,9.27) | 1.62 (-3.59,6.84) |

BMI, body mass index; CI, confidence interval; yr, years

^a^ BMI is presented in the natural log and values represent the mean predicted natural log of BMI at each age shown.

^b^ Differences at each age are back transformed from the log scale and are interpreted as the percentage difference in the mean level in original units at each age comparing each category with the reference trajectory.

**Table S13:** Mean trajectories of fat mass and mean differences by haplogroup, estimated from multilevel models estimated from multilevel models

|  | **Mean log fat mass trajectory (95% CI) in haplogroup H (kg or kg/yr** **) (reference)^a^** | **Mean difference in fat mass trajectory (95% CI) comparing with haplogroup H(% or %/yr)^b^** | | | | | | | |
| --- | --- | --- | --- | --- | --- | --- | --- | --- | --- |
|  |  | **Haplogroup U** | **Haplogroup T** | **Haplogroup J** | **Haplogroup K** | **Haplogroup V** | **Haplogroup W** | **Haplogroup I** | **Haplogroup X** |
| **Female** |  |  |  |  |  |  |  |  |  |
| Age 9yr (kg) or (%) | 2.02 (1.99,2.05) | -0.82 (-6.56,4.93) | 1.12 (-5.31,7.56) | 1.09 (-5.25,7.42) | -1.77 (-8.88,5.34) | -9.33 (-16.71,-1.94) | -12.37 (-25.51,0.77) | 2.70 (-8.60,13.99) | -16.39 (-29.26,-3.51) |
| Change 9-13yr (kg/yr) or (%/yr) | 0.15 (0.15,0.16) | -0.06 (-1.14,1.03) | -0.54 (-1.72,0.64) | 0.08 (-1.10,1.27) | -0.76 (-2.10,0.58) | -0.23 (-1.75,1.30) | 0.99 (-1.87,3.85) | -0.81 (-2.84,1.22) | 0.94 (-1.94,3.83) |
| Change 13-15yr (kg/yr) or (%/yr) | 0.10 (0.09,0.11) | 0.56 (-1.42,2.53) | -0.78 (-2.93,1.37) | -0.21 (-2.34,1.92) | 0.82 (-1.67,3.31) | 1.70 (-1.15,4.55) | 3.10 (-2.09,8.28) | 0.78 (-2.94,4.49) | 3.81 (-1.79,9.41) |
| Change 15-18yr (kg/yr) or (%/yr) | 0.06 (0.05,0.06) | -0.99 (-2.37,0.38) | -0.32 (-1.77,1.14) | 0.11 (-1.38,1.59) | 0.29 (-1.39,1.97) | 1.86 (-0.15,3.86) | -0.46 (-4.06,3.14) | -0.13 (-2.75,2.49) | 1.01 (-2.81,4.83) |
| Age 18yr (kg) or (%) | 3.01 (2.99,3.04) | -2.88 (-7.74,1.97) | -3.49 (-8.66,1.68) | 1.21 (-6.83,9.24) | -4.40 (-13.11,4.31) | 0.40 (-10.03,10.84) | 6.70 (-13.74,27.14) | -1.95 (-15.60,11.70) | 8.90 (-12.76,30.55) |
| **Male** |  |  |  |  |  |  |  |  |  |
| Age 9yr (kg) or (%) | 1.70 (1.67,1.73) | 1.67 (-5.48,8.83) | -4.63 (-11.92,2.67) | 2.70 (-5.08,10.48) | -0.41 (-8.40,7.58) | -2.81 (-13.06,7.45) | 1.49 (-15.06,18.03) | 10.76 (-5.08,26.59) | -10.46 (-26.73,5.81) |
| Change 9-13yr (kg/yr) or (%/yr) | 0.15 (0.15,0.16) | -0.02 (-1.56,1.52) | 0.08 (-1.60,1.77) | -1.01 (-2.68,0.67) | 0.65 (-1.11,2.41) | 0.76 (-1.63,3.15) | 1.05 (-2.63,4.73) | 1.61 (-1.47,4.70) | 3.57 (-0.44,7.58) |
| Change 13-15yr (kg/yr) or (%/yr) | -0.08 (-0.09,-0.06) | -0.65 (-3.48,2.18) | 0.85 (-2.30,4.00) | -0.62 (-3.69,2.45) | -2.92 (-6.00,0.15) | -3.73 (-8.03,0.58) | 4.18 (-2.91,11.26) | 3.44 (-2.13,9.01) | 2.48 (-4.30,9.27) |
| Change 15-18yr (kg/yr) or (%/yr) | 0.10 (0.09,0.11) | -0.40 (-2.70,1.90) | 2.48 (-0.10,5.07) | 0.76 (-1.74,3.26) | 1.60 (-1.02,4.21) | 1.43 (-2.14,5.01) | -4.18 (-9.34,0.97) | -3.45 (-7.88,0.98) | 0.55 (-5.05,6.15) |
| Age 18yr (kg) or (%) | 2.46 (2.42,2.50) | -0.92 (-9.24,7.41) | 4.75 (-4.98,14.49) | -7.16 (-19.31,4.99) | -1.72 (-15.28,11.85) | -6.71 (-23.66,10.24) | 15.50 (-18.15,49.14) | 31.80 (0.28,63.32) | 37.72 (-4.25,79.68) |

CI, confidence interval; kg/yr, kilograms per year; %/yr, percentage per year.

^a^Fat mass was transformed using the natural log. All predicted mean values (kg) and rates of change per year (kg/yr) are on the log scale

^b^The difference between haplogroups is back transformed from the log scale for ease of interpretation and is interpreted as the percentage difference in the mean level comparing each category with haplogroup H or percentage difference in change per year (%/yr) comparing each category with haplogroup H.

**Table S14:** Mean trajectories of lean mass and mean differences by haplogroup, estimated from multilevel models

|  | **Mean trajectory (95% CI) in haplogroup H (reference)** | **Mean difference in trajectory (95% CI) comparing with haplogroup H** | | | | | | | |
| --- | --- | --- | --- | --- | --- | --- | --- | --- | --- |
|  |  | **Haplogroup U** | **Haplogroup T** | **Haplogroup J** | **Haplogroup K** | **Haplogroup V** | **Haplogroup W** | **Haplogroup I** | **Haplogroup X** |
| **Female** |  |  |  |  |  |  |  |  |  |
| Age 9yr (kg) | 20.92 (20.76,21.07) | -0.25 (-0.57,0.08) | -0.21 (-0.57,0.15) | -0.004 (-0.36,0.35) | -0.37 (-0.77,0.03) | -0.39 (-0.85,0.06) | -0.99 (-1.84,-0.14) | 0.14 (-0.48,0.76) | -1.34 (-2.20,-0.47) |
| Change 9-13yr (kg/yr) | 3.17 (3.12,3.22) | 0.02 (-0.08,0.12) | 0.10 (-0.01,0.21) | 0.09 (-0.03,0.20) | 0.01 (-0.117,0.14) | -0.09 (-0.24,0.05) | 0.18 (-0.09,0.45) | -0.07 (-0.27,0.12) | -0.03 (-0.30,0.25) |
| Change 13-15yr (kg/yr) | 1.72 (1.61,1.83) | 0.18 (-0.05,0.41) | -0.09 (-0.34,0.16) | -0.11 (-0.36,0.14) | 0.07 (-0.22,0.35) | 0.17 (-0.16,0.50) | -0.04 (-0.62,0.55) | 0.44 (0.003,0.87) | -0.01 (-0.64,0.62) |
| Change 15-18yr (kg/yr) | 0.40 (0.35,0.46) | -0.07 (-0.18,0.04) | -0.04 (-0.16,0.08) | -0.04 (-0.16,0.08) | 0.01 (-0.13,0.14) | -0.18 (-0.34,-0.02) | 0.13 (-0.16,0.41) | -0.19 (-0.40,0.02) | 0.02 (-0.29,0.33) |
| Age 18yr (kg) | 38.25 (38.01,38.49) | -0.01 (-0.52,0.49) | -0.10 (-0.65,0.44) | 0.01 (-0.54,0.56) | -0.16 (-0.78,0.46) | -0.97 (-1.68,-0.25) | 0.04 (-1.30,1.37) | 0.15 (-0.81,1.11) | -1.41 (-2.80,-0.02) |
| **Male** |  |  |  |  |  |  |  |  |  |
| Age 9yr (kg) | 22.88 (22.72,23.04) | 0.06 (-0.27,0.40) | 0.02 (-0.34,0.39) | -0.13 (-0.49,0.23) | 0.20 (-0.18,0.59) | -0.45 (-0.97,0.07) | -0.05 (-0.84,0.74) | 0.11 (-0.57,0.79) | 0.53 (-0.33,1.38) |
| Change 9-13yr (kg/yr) | 2.79 (2.70,2.87) | 0.09 (-0.09,0.27) | 0.17 (-0.03,0.36) | 0.08 (-0.11,0.28) | 0.09 (-0.12,0.29) | -0.01 (-0.29,0.26) | 0.09 (-0.35,0.52) | 0.26 (-0.09,0.61) | -0.19 (-0.64,0.27) |
| Change 13-15yr (kg/yr) | 7.34 (7.20,7.48) | -0.15 (-0.46,0.15) | -0.03 (-0.36,0.30) | -0.005 (-0.33,0.32) | 0.23 (-0.10,0.57) | 0.15 (-0.33,0.63) | -0.62 (-1.34,0.11) | -0.13 (-0.70,0.44) | 0.13 (-0.57,0.83) |
| Change 15-18yr (kg/yr) | 2.25 (2.12,2.38) | 0.01 (-0.26,0.29) | -0.14 (-0.43,0.16) | -0.13 (-0.43,0.16) | -0.17 (-0.47,0.14) | 0.0003 (-0.41,0.41) | 0.15 (-0.48,0.79) | -0.05 (-0.59,0.49) | -0.45 (-1.12,0.21) |
| Age 18yr (kg) | 55.45 (55.05,55.84) | 0.15 (-0.68,0.98) | 0.21 (-0.70,1.12) | -0.22 (-1.11,0.68) | 0.52 (-0.42,1.45) | -0.19 (-1.42,1.04) | -0.47 (-2.42,1.47) | 0.74 (-0.93,2.40) | -1.32 (-3.39,0.75) |

kg/yr, kilograms per year; yr

**Table S15:** Mean trajectories of SBP, DBP and pulse rate and mean differences by haplogroup, estimated from multilevel models

|  | **Mean trajectory (95% CI) in haplogroup H (reference)^a^** | **Mean difference in trajectory (95% CI) comparing with haplogroup H^a^** | | | | | | | |
| --- | --- | --- | --- | --- | --- | --- | --- | --- | --- |
|  |  | **Haplogroup U** | **Haplogroup T** | **haplogroup J** | **haplogroup K** | **Haplogroup V** | **Haplogroup W** | **Haplogroup I** | **Haplogroup X** |
| **SBP**  **Females** |  |  |  |  |  |  |  |  |  |
| Age 7yr (mmHg) | 98.32 (97.81,98.83) | -0.37 (-1.44,0.69) | -0.73 (-1.90,0.44) | -0.57 (-1.73,0.60) | -0.47 (-1.78,0.85) | -1.18 (-2.68,0.33) | 0.27 (-2.57,3.10) | -0.36 (-2.38,1.65) | -3.36 (-6.19,-0.53) |
| Change 7-12yr (mmHg/yr) | 1.82 (1.70,1.94) | 0.07 (-0.18,0.32) | 0.20 (-0.07,0.48) | 0.10 (-0.18,0.37) | -0.12 (-0.44,0.19) | 0.02 (-0.34,0.37) | -0.39 (-1.05,0.27) | -0.12 (-0.60,0.36) | 0.64 (-0.02,1.31) |
| Change 12-16yr (mmHg/yr) | 3.89 (3.69,4.09) | -0.23 (-0.64,0.19) | -0.36 (-0.80,0.09) | -0.07 (-0.52,0.37) | 0.27 (-0.25,0.79) | 0.24 (-0.34,0.82) | 0.66 (-0.42,1.75) | 0.27 (-0.52,1.05) | -0.32 (-1.45,0.82) |
| Change 16-18yr (mmHg/yr) | -6.04 (-6.48,-5.61) | 0.53 (-0.37,1.44) | 0.64 (-0.31,1.59) | 0.30 (-0.69,1.28) | 0.08 (-1.07,1.22) | -0.02 (-1.30,1.26) | 0.85 (-1.50,3.21) | 0.16 (-1.53,1.85) | -0.24 (-2.69,2.20) |
| Age 18yr (mmHg) | 110.91 (110.32,111.49) | 0.12 (-1.08,1.32) | 0.15 (-1.12,1.41) | 0.22 (-1.10,1.55) | 0.14 (-1.36,1.63) | -0.17 (-1.94,1.59) | 2.68 (-0.50,5.85) | 0.40 (-1.92,2.73) | -1.89 (-5.21,1.42) |
| **Males** |  |  |  |  |  |  |  |  |  |
| Age 7yr (mmHg) | 97.96 (97.47,98.45) | 0.07 (-0.96,1.11) | -0.63 (-1.75,0.49) | -0.37 (-1.48,0.73) | 0.19 (-0.98,1.36) | 0.45 (-1.11,2.00) | -0.74 (-3.02,1.54) | -0.24 (-2.30,1.83) | -1.96 (-4.63,0.70) |
| Change 7-12yr (mmHg/yr) | 1.58 (1.46,1.70) | 0.12 (-0.13,0.37) | 0.18 (-0.09,0.44) | 0.12 (-0.14,0.39) | -0.05 (-0.32,0.23) | -0.19 (-0.55,0.18) | -0.06 (-0.61,0.49) | 0.83 (0.34,1.32) | 0.58 (-0.05,1.21) |
| Change 12-16yr (mmHg/yr) | 5.82 (5.62,6.02) | -0.16 (-0.57,0.26) | -0.16 (-0.61,0.29) | 0.23 (-0.21,0.68) | 0.23 (-0.22,0.68) | 0.37 (-0.26,1.00) | -0.10 (-1.10,0.90) | -1.23 (-2.02,-0.44) | -0.47 (-1.46,0.51) |
| Change 16-18yr (mmHg/yr) | -3.72 (-4.20,-3.24) | -0.07 (-1.07,0.92) | 0.20 (-0.89,1.28) | -0.58 (-1.65,0.49) | 0.23 (-0.84,1.31) | -1.22 (-2.70,0.26) | -1.05 (-3.37,1.26) | 0.45 (-1.57,2.47) | 0.37 (-1.99,2.73) |
| Age 18yr (mmHg) | 121.68 (120.96,122.40) | -0.10 (-1.59,1.40) | 0.01 (-1.61,1.64) | 0.02 (-1.58,1.62) | 1.34 (-0.31,2.99) | -1.44 (-3.60,0.72) | -3.54 (-6.93,-0.15) | -0.12 (-3.19,2.96) | -0.23 (-3.91,3.46) |
| **DBP**  **Females** |  |  |  |  |  |  |  |  |  |
| Age 7yr (mmHg) | 57.17 (56.82,57.53) | -0.50 (-1.25,0.25) | -0.32 (-1.14,0.50) | -0.23 (-1.05,0.59) | -0.67 (-1.60,0.25) | -0.25 (-1.31,0.81) | -1.74 (-3.74,0.26) | 0.41 (-1.01,1.83) | -1.49 (-3.48,0.50) |
| Change 7-12yr (mmHg/yr) | 0.07 (-0.03,0.16) | 0.16 (-0.03,0.35) | 0.09 (-0.12,0.30) | -0.01 (-0.22,0.20) | 0.04 (-0.19,0.28) | -0.01 (-0.28,0.25) | 0.34 (-0.16,0.83) | -0.15 (-0.51,0.21) | 0.24 (-0.27,0.74) |
| Change 12-16yr (mmHg/yr) | 2.45 (2.29,2.62) | -0.20 (-0.54,0.14) | -0.26 (-0.62,0.10) | 0.19 (-0.18,0.55) | -0.20 (-0.63,0.22) | -0.18 (-0.65,0.29) | -0.58 (-1.47,0.32) | -0.16 (-0.80,0.48) | -0.14 (-1.07,0.79) |
| Change 16-18yr (mmHg/yr) | -1.30 (-1.66,-0.95) | 0.27 (-0.47,1.02) | 0.45 (-0.32,1.23) | -0.60 (-1.41,0.22) | 0.31 (-0.63,1.25) | 0.76 (-0.28,1.80) | 1.88 (-0.07,3.84) | 0.52 (-0.84,1.88) | 0.52 (-1.48,2.52) |
| Age 18yr (mmHg) | 64.72 (64.29,65.15) | 0.05 (-0.83,0.93) | -0.01 (-0.93,0.90) | -0.74 (-1.71,0.24) | -0.65 (-1.75,0.45) | 0.47 (-0.81,1.76) | 1.40 (-0.93,3.72) | 0.06 (-1.61,1.73) | 0.18 (-2.25,2.61) |
| **Males** |  |  |  |  |  |  |  |  |  |
| Age 7yr (mmHg) | 56.12 (55.76,56.49) | 0.28 (-0.49,1.05) | 0.03 (-0.80,0.86) | 0.22 (-0.60,1.04) | -0.13 (-1.00,0.74) | -0.08 (-1.24,1.07) | 0.12 (-1.57,1.81) | 0.54 (-0.99,2.08) | -0.66 (-2.63,1.32) |
| Change 7-12yr (mmHg/yr) | 0.14 (0.04,0.23) | 0.05 (-0.15,0.24) | 0.02 (-0.19,0.23) | 0.02 (-0.19,0.23) | -0.07 (-0.29,0.16) | 0.17 (-0.12,0.46) | -0.01 (-0.45,0.43) | 0.20 (-0.19,0.59) | 0.05 (-0.45,0.55) |
| Change 12-16yr (mmHg/yr) | 2.88 (2.69,3.07) | 0.13 (-0.27,0.52) | -0.003 (-0.43,0.43) | -0.10 (-0.52,0.33) | 0.05 (-0.38,0.48) | -0.28 (-0.88,0.32) | -0.46 (-1.41,0.50) | -0.30 (-1.06,0.46) | 0.37 (-0.57,1.31) |
| Change 16-18yr (mmHg/yr) | -2.65 (-3.09,-2.22) | -0.62 (-1.53,0.28) | 0.07 (-0.92,1.05) | 0.10 (-0.88,1.07) | 0.58 (-0.39,1.56) | 0.01 (-1.33,1.35) | 1.08 (-1.05,3.21) | -0.36 (-2.23,1.52) | -0.19 (-2.34,1.97) |
| Age 18yr (mmHg) | 63.03 (62.52,63.55) | -0.22 (-1.28,0.85) | 0.26 (-0.90,1.43) | 0.15 (-1.00,1.29) | 0.91 (-0.25,2.07) | -0.34 (-1.86,1.18) | 0.39 (-2.01,2.79) | -0.35 (-2.66,1.95) | 0.70 (-1.94,3.33) |
| **Pulse rate**  **Females** |  |  |  |  |  |  |  |  |  |
| Age 7yr (bpm) | 85.72 (85.14,86.30) | -0.33 (-1.55,0.88) | 0.31 (-1.03,1.64) | 0.79 (-0.54,2.12) | 0.92 (-0.58,2.42) | -0.50 (-2.22,1.22) | 2.32 (-0.93,5.56) | 0.64 (-1.66,2.94) | -0.43 (-3.64,2.79) |
| Change 7-12yr (bpm/yr) | -1.79 (-1.93,-1.65) | 0.18 (-0.11,0.46) | -0.07 (-0.38,0.25) | -0.11 (-0.42,0.21) | 0.04 (-0.32,0.40) | 0.11 (-0.30,0.51) | -0.55 (-1.31,0.20) | -0.18 (-0.73,0.37) | -0.09 (-0.85,0.67) |
| Change 12-16yr (bpm/yr) | -0.27 (-0.48,-0.05) | 0.020 (-0.42,0.46) | -0.15 (-0.62,0.32) | 0.01 (-0.46,0.49) | -0.40 (-0.95,0.15) | 0.02 (-0.59,0.64) | 0.25 (-0.90,1.40) | -0.06 (-0.89,0.77) | 0.86 (-0.34,2.07) |
| Change 16-18yr (bpm/yr) | -4.76 (-5.22,-4.29) | 0.49 (-0.48,1.46) | 0.30 (-0.71,1.31) | -0.75 (-1.80,0.31) | 0.43 (-0.80,1.66) | 0.58 (-0.80,1.95) | 0.05 (-2.47,2.57) | -0.22 (-2.02,1.58) | -1.97 (-4.60,0.67) |
| Age 18yr (bpm) | 66.19 (65.51,66.87) | 1.63 (0.23,3.03) | -0.03 (-1.50,1.44) | -1.18 (-2.73,0.36) | 0.35 (-1.39,2.09) | 1.28 (-0.77,3.33) | 0.64 (-3.07,4.36) | -0.95 (-3.64,1.75) | -1.35 (-5.22,2.51) |
| **Males** |  |  |  |  |  |  |  |  |  |
| Age 7yr (bpm) | 83.02 (82.45,83.59) | -0.05 (-1.25,1.16) | -0.92 (-2.22,0.39) | 0.54 (-0.75,1.83) | -0.16 (-1.52,1.20) | 1.09 (-0.72,2.91) | 0.51 (-2.15,3.16) | 0.50 (-1.91,2.92) | -3.16 (-6.27,-0.05) |
| Change 7-12yr (bpm/yr) | -1.91 (-2.05,-1.76) | -0.04 (-0.34,0.25) | 0.30 (-0.02,0.62) | 0.07 (-0.25,0.39) | 0.004 (-0.33,0.34) | 0.12 (-0.32,0.56) | -0.42 (-1.09,0.25) | 0.17 (-0.42,0.77) | 0.67 (-0.10,1.43) |
| Change 12-16yr (bpm/yr) | -0.75 (-0.97,-0.54) | -0.02 (-0.47,0.44) | -0.38 (-0.87,0.12) | 0.06 (-0.44,0.55) | -0.22 (-0.72,0.28) | -0.30 (-0.99,0.40) | 0.56 (-0.54,1.66) | 0.14 (-0.73,1.01) | -0.11 (-1.20,0.99) |
| Change 16-18yr (bpm/yr) | -4.16 (-4.67,-3.66) | -0.12 (-1.17,0.93) | 0.06 (-1.09,1.21) | -0.34 (-1.47,0.79) | 0.29 (-0.84,1.42) | -0.41 (-1.96,1.14) | -0.48 (-2.93,1.96) | -1.20 (-3.37,0.97) | 0.03 (-2.48,2.54) |
| Age 18yr (bpm) | 62.15 (61.41,62.88) | -0.58 (-2.10,0.94) | -0.81 (-2.47,0.85) | 0.42 (-1.21,2.04) | -0.44 (-2.11,1.23) | -0.31 (-2.48,1.87) | -0.31 (-3.76,3.13) | -0.48 (-3.67,2.71) | -0.19 (-3.94,3.57) |

DBP, diastolic blood pressure; SBP, systolic blood pressure; CI, confidence interval

^a^Units are presented in mmHg for SBP and DBP and bpm for pulse rate

**Table S16:** Mean trajectories of HDL-c and non-HDL-c and mean differences by haplogroup, estimated from multilevel models estimated from multilevel models

|  | **Mean trajectory (95% CI) in haplogroup H (reference)** | **Mean difference in trajectory (95% CI) comparing with haplogroup H** | | | | | | | |
| --- | --- | --- | --- | --- | --- | --- | --- | --- | --- |
|  |  | **Haplogroup U** | **Haplogroup T** | **Haplogroup J** | **Haplogroup K** | **Haplogroup V** | **Haplogroup W** | **Haplogroup I** | **Haplogroup X** |
| **HDL-c**  **Females** |  |  |  |  |  |  |  |  |  |
| Birth (mmol/l) | 0.79 (0.49,1.09) | -0.24 (-0.87,0.39) | 0.12 (-0.56,0.81) | 0.12 (-0.54,0.78) | 0.13 (-0.69,0.95) | 0.37 (-0.48,1.22) | -0.23 (-1.82,1.35) | 1.23 (-0.10,2.56) | -0.25 (-2.06,1.56) |
| Change 0-7yr (mmol/l/yr) | 0.09 (0.05,0.14) | 0.04 (-0.05,0.13) | -0.02 (-0.12,0.08) | -0.02 (-0.11,0.08) | -0.01 (-0.13,0.10) | -0.05 (-0.17,0.08) | 0.04 (-0.19,0.26) | -0.18 (-0.37,0.01) | 0.03 (-0.23,0.29) |
| Change 7-18yr (mmol/l/yr) | -0.01 (-0.01,-0.01) | 0.0003 (-0.004,0.004) | 0.0003 (-0.004,0.01) | -0.005 (-0.01,-0.0003) | -0.006 (-0.01,-0.002) | 0.0002 (-0.01,0.006) | -0.01 (-0.02,0.003) | -0.004 (-0.01,0.004) | 0.005 (-0.01,0.02) |
| Age 18yr (mmol/l) | 1.33 (1.31,1.35) | 0.02 (-0.02,0.06) | -0.003 (-0.05,0.04) | -0.04 (-0.09,0.002) | -0.04 (-0.09,0.004) | 0.05 (-0.003,0.11) | -0.07 (-0.18,0.05) | -0.05 (-0.13,0.03) | 0.02 (-0.09,0.13) |
| **Males** |  |  |  |  |  |  |  |  |  |
| Birth (mmol/l) | 0.81 (0.59,1.03) | -0.25 (-0.72,0.21) | -0.33 (-0.85,0.19) | -0.32 (-0.82,0.19) | -0.31 (-0.85,0.23) | -0.34 (-1.00,0.31) | 2.13 (1.08,3.18) | -0.30 (-1.33,0.73) | -0.38 (-1.64,0.87) |
| Change 0-7yr (mmol/l/yr) | 0.10 (0.07,0.14) | 0.03 (-0.03,0.10) | 0.05 (-0.02,0.12) | 0.04 (-0.03,0.11) | 0.05 (-0.03,0.13) | 0.05 (-0.05,0.14) | -0.30 (-0.45,-0.15) | 0.05 (-0.10,0.20) | 0.05 (-0.13,0.23) |
| Change 7-18yr (mmol/l/yr) | -0.04 (-0.04,-0.04) | 0.003 (-0.001,0.01) | 0.001 (-0.003,0.005) | 0.003 (-0.001,0.01) | -0.0004 (-0.01,0.004) | 0.001 (-0.005,0.007) | 0.004 (-0.01,0.01) | -0.004 (-0.01,0.003) | 0.003 (-0.01,0.01) |
| Age 18yr (mmol/l) | 1.14 (1.12,1.15) | 0.02 (-0.02,0.05) | 0.04 (-0.003,0.08) | 0.01 (-0.03,0.05) | 0.03 (-0.008,0.07) | -0.01 (-0.07,0.04) | 0.05 (-0.03,0.14) | 0.01 (-0.06,0.08) | -0.01 (-0.10,0.08) |
| **Non-HDL-c**  **Females** |  |  |  |  |  |  |  |  |  |
| Birth (mmol/l) | 1.68 (1.44,1.91) | -0.04 (-0.53,0.45) | -0.12 (-0.65,0.42) | -0.16 (-0.68,0.36) | 0.24 (-0.38,0.85) | -0.42 (-1.09,0.26) | 0.58 (-0.67,1.83) | -0.47 (-1.45,0.51) | 0.29 (-1.10,1.67) |
| Change 0-9yr (mmol/l/yr) | 0.16 (0.14,0.19) | 0.003 (-0.06,0.06) | 0.01 (-0.05,0.08) | 0.009 (-0.06,0.07) | -0.03 (-0.11,0.04) | 0.04 (-0.05,0.12) | -0.06 (-0.22,0.09) | 0.05 (-0.07,0.17) | -0.04 (-0.21,0.13) |
| Change 9-18yr (mmol/l/yr) | -0.07 (-0.08,-0.07) | 0.005 (-0.01,0.02) | -0.004 (-0.02,0.01) | 0.003 (-0.01,0.02) | 0.01 (-0.01,0.03) | 0.01 (-0.01,0.03) | 0.01 (-0.03,0.05) | -0.01 (-0.041,0.01) | 0.04 (0.003,0.08) |
| Age 18yr (mmol/l) | 2.50 (2.45,2.54) | 0.03 (-0.06,0.12) | -0.05 (-0.14,0.05) | -0.06 (-0.16,0.04) | 0.04 (-0.08,0.15) | -0.001 (-0.13,0.13) | 0.11 (-0.15,0.38) | -0.13 (-0.32,0.05) | 0.28 (0.03,0.53) |
| **Males** |  |  |  |  |  |  |  |  |  |
| Birth (mmol/l) | 1.40 (1.22,1.58) | 0.23 (-0.15,0.60) | 0.29 (-0.13,0.70) | 0.22 (-0.19,0.62) | 0.29 (-0.15,0.72) | 0.14 (-0.41,0.69) | -1.54 (-2.41,-0.67) | 0.30 (-0.52,1.11) | 0.11 (-0.86,1.09) |
| Change 0-9yr (mmol/l/yr) | 0.17 (0.15,0.19) | -0.03 (-0.07,0.02) | -0.03 (-0.09,0.02) | -0.03 (-0.08,0.02) | -0.03 (-0.08,0.02) | -0.01 (-0.08,0.06) | 0.19 (0.09,0.30) | -0.035 (-0.13,0.06) | -0.005 (-0.12,0.11) |
| Change 9-18yr (mmol/l/yr) | -0.07 (-0.08,-0.07) | 0.01 (-0.005,0.02) | 0.01 (0.000,0.02) | 0.01 (-0.002,0.02) | 0.01 (-0.005,0.02) | -0.002 (-0.02,0.01) | -0.003 (-0.03,0.02) | 0.0016 (-0.02,0.02) | -0.001 (-0.03,0.03) |
| Age 18yr (mmol/l) | 2.27 (2.23,2.31) | 0.03 (-0.06,0.11) | 0.07 (-0.02,0.17) | 0.05 (-0.04,0.14) | 0.10 (0.01,0.20) | 0.024 (-0.11,0.16) | 0.16 (-0.06,0.38) | -0.002 (-0.17,0.16) | 0.06 (-0.14,0.27) |

CI, confidence interval; HDL-c, high density lipoprotein cholesterol; mmol/l, millimole per litre; mmol/l, millimole per litre per year

**Table S17:** Mean trajectories of triglyceride estimated from multilevel models**,** by haplogroup

|  | **Mean trajectory (95% CI) in haplogroup H (reference)^a^** | **Mean difference in trajectory (95% CI) comparing with haplogroup H^b^** | | | | | | | |
| --- | --- | --- | --- | --- | --- | --- | --- | --- | --- |
|  |  | **Haplogroup U** | **Haplogroup T** | **Haplogroup J** | **Haplogroup K** | **Haplogroup V** | **Haplogroup W** | **Haplogroup I** | **Haplogroup X** |
| **Female** |  |  |  |  |  |  |  |  |  |
| Birth (mmol/l or %) | -0.67 (-0.70,-0.64) | 1.20 (-5.81,8.21) | -1.52 (-8.94,5.91) | 0.88 (-6.50,8.26) | -0.58 (-9.52,8.36) | -5.67 (-14.57,3.23) | 10.05 (-9.31,29.41) | -2.67 (-16.79,11.45) | 9.84 (-12.14,31.82) |
| Change 0-9yr (mmol/l/yr or %/yr) | 0.09 (0.08,0.09) | 0.03 (-0.96,1.02) | 0.52 (-0.57,1.61) | -0.25 (-1.30,0.80) | -0.43 (-1.69,0.83) | 0.45 (-0.92,1.83) | -1.70 (-4.16,0.76) | 0.59 (-1.47,2.64) | -1.32 (-4.07,1.42) |
| Change 9-18yr (mmol/l/yr or %/yr) | -0.05 (-0.05,-0.04) | 0.07 (-0.69,0.82) | -0.17 (-0.96,0.63) | 0.63 (-0.18,1.44) | 0.88 (-0.02,1.78) | 0.74 (-0.36,1.83) | 1.29 (-0.85,3.43) | -0.03 (-1.50,1.44) | 1.06 (-1.00,3.13) |
| Age 18yr (mmol/l or %) | -0.31 (-0.34,-0.29) | 2.09 (-3.18,7.36) | 1.61 (-3.83,7.04) | 4.36 (-1.41,10.13) | 3.42 (-2.86,9.69) | 4.94 (-2.69,12.57) | 5.84 (-9.95,21.64) | 2.30 (-7.98,12.58) | 7.13 (-8.05,22.31) |
| **Male** |  |  |  |  |  |  |  |  |  |
| Birth (mmol/l or %) | -0.70 (-0.73,-0.66) | 2.58 (-4.45,9.62) | 1.96 (-5.89,9.81) | 5.04 (-2.81,12.89) | 1.89 (-6.28,10.06) | 6.39 (-4.00,16.77) | 7.48 (-9.26,24.22) | 9.11 (-7.58,25.79) | -1.56 (-20.24,17.13) |
| Change 0-9yr (mmol/l/yr or %/yr) | 0.08 (0.08,0.08) | -0.34 (-1.33,0.64) | 0.03 (-1.07,1.13) | -0.81 (-1.86,0.25) | -0.25 (-1.39,0.89) | -0.15 (-1.57,1.27) | -1.17 (-3.39,1.05) | -0.87 (-2.97,1.24) | 0.22 (-2.45,2.88) |
| Change 9-18yr (mmol/l/yr or %/yr) | -0.04 (-0.04,-0.04) | -0.28 (-1.11,0.55) | -0.18 (-1.07,0.72) | 0.20 (-0.69,1.08) | 0.44 (-0.48,1.37) | 0.20 (-1.08,1.48) | 1.50 (-0.53,3.53) | -0.32 (-1.91,1.28) | 0.61 (-1.44,2.67) |
| Age 18yr (mmol/l or %) | -0.33 (-0.36,-0.30) | -3.02 (-8.57,2.53) | 0.62 (-5.68,6.91) | -0.61 (-6.67,5.44) | 3.68 (-2.90,10.26) | 6.83 (-2.70,16.36) | 10.55 (-5.56,26.65) | -1.93 (-12.57,8.72) | 6.06 (-8.58,20.71) |

CI, confidence interval; mmol/l, millimole per litre; mmol/l/year, millimoles per litre per year; %/yr, percentage per year

^a^Triglyceride was transformed using the natural log. All predicted mean values (mmol/l) and rates of change per year (mmol/l/yr) are on the log scale

^b^The difference between haplogroups is back transformed from the log scale for ease of interpretation and is interpreted as the percentage difference in the mean level comparing each category with haplogroup H or percentage difference in change per year (%/yr) comparing each category with haplogroup H
